# PCSK9 and breast cancer survival: a Mendelian Randomization study

**DOI:** 10.1101/2025.10.03.25337270

**Authors:** Janne Pott, Amy Mason, Ville Salo, FINNGEN, Johannes Kettunen, Stephen Burgess

## Abstract

**Background:** Proprotein convertase subtilisin/kexin type 9 (PCSK9) is well known for its causal effects on the lipid metabolism. A recent study identified an association between rs562556 within *PCSK9* and breast cancer survival (BCS). It was suggested that PCSK9 inhibition might allow for early intervention strategies to prevent metastasizing breast cancer. Here, we attempt to replicate these findings by using genome-wide association study (GWAS) summary statistics in three 2-sample Mendelian Randomization (MR) approaches.

**Methods:** We used publicly available GWAS summary statistics for PCSK9 and BCS from the Breast Cancer Association Consortium (N=99,217), and performed SNP association tests with BCS in the FinnGen study (N=4,648). First, we tested the MR-ratio using only the reported SNP rs562556, then tested the genetically proxied PCSK9 inhibition using the MR inverse variance weighted (IVW) approach, and finally adjusted for indirect effects of the lipid metabolism using multivariable MR (MVMR-IVW). Coronary artery disease and parental longevity were chosen as positive control outcomes.

**Results:** We found no significant association between PCSK9 and BCS in neither the MR-ratio, MR-IVW nor MVMR-IVW approach. The positive controls were significant throughout.

**Conclusion:** A significant positive effect of PCSK9 on BCS could only be reproduced when using the outcome data from the recent study, but not when using independent, larger GWASs.

**Impact:** Potential reasons for the discrepant results are different genetic models, sample selection criteria, and the time-variability of Hazard Ratio estimates. They should be explored before considering PCSK9 a therapeutic target in BC patients.

## Introduction

Proprotein convertase subtilisin/kexin type 9 (PCSK9) is mainly expressed in the liver and plays a key role in lipid metabolism [1]. It binds to low-density lipoprotein receptors (LDL-R) on the surface of hepatocytes, and prohibits LDL-R recycling after its internalization [2]. Hence, PCSK9 leads to degradation of LDL-R and reduction of LDL cholesterol (LDL-C) clearance in the blood flow (see Figure 1A). PCSK9’s pivotal role in LDL-C clearance has led to development of various PCSK9 inhibitors to treat hypercholesterolemia [3–5], which are usually given when other lipid-lowering medication such as statins are not tolerated or fail to hit the target reduction of LDL-C levels.

**Figure 1:**
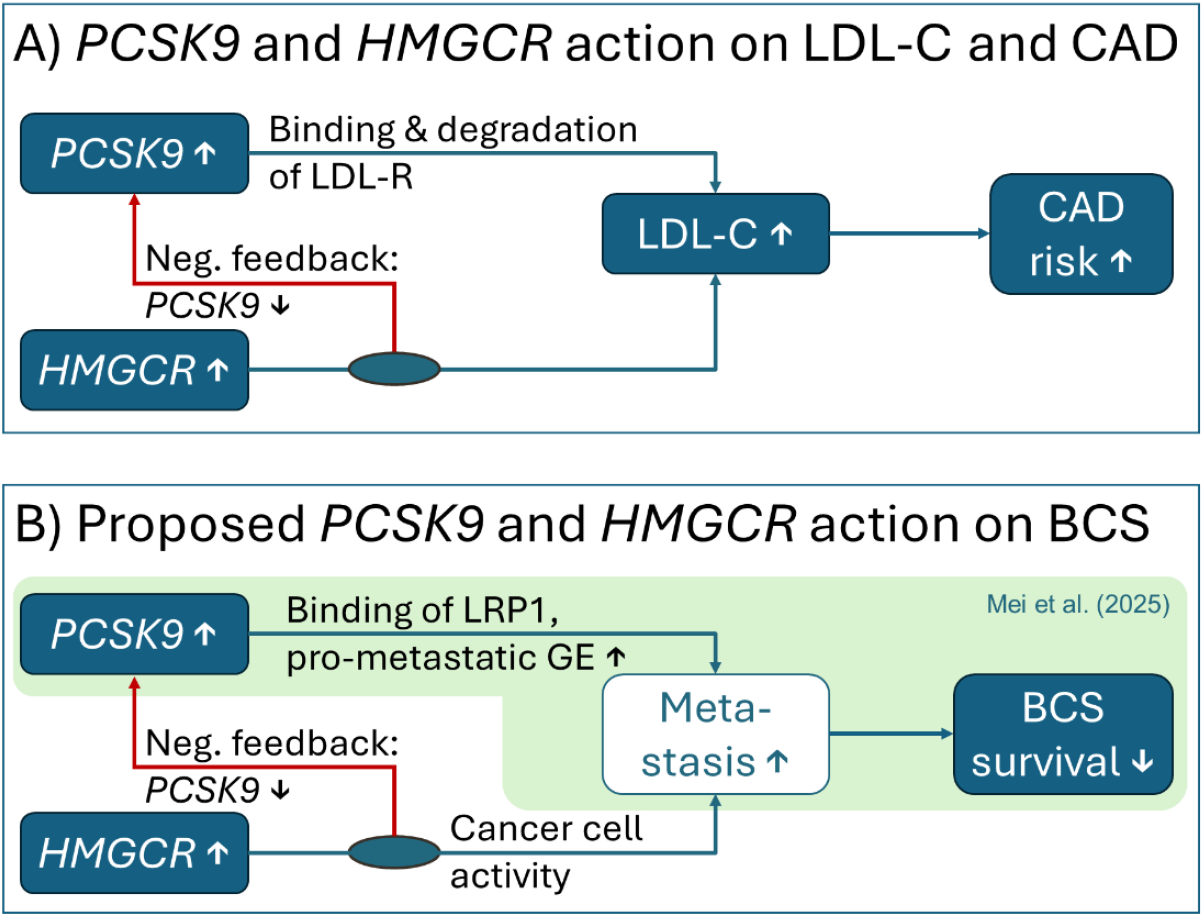
Directed acyclic graphs (DAGs). Blue filled boxes indicate considered exposures and outcomes in this study. A) Classic path of action for PCSK9 and LDL-C levels on risk for coronary artery disease. PCSK9 binds to LDL-receptors (LDLR) and leads to its degradation, resulting in higher LDL-C levels. HMG-CoA activity activates the intracellular cholesterol biosynthesis, resulting in reduced LDLR expression. LDLR expression is regulated by a shared transcription factor of PCSK9. Hence PCSK9 levels are also reduced. B) Considered path of action of PCSK9 on breast cancer survival (BCS) in this work. Mei et al. *[14]* reported higher levels of PCSK9 associated with breast cancer survival, presumable due to its binding to LDL-R related protein 1 (LRP1), which in turn leads to increased gene expression (GE) of pro-metastatic genes. The induces metastases are then the cause for the increased mortality. It is yet unclear how HMG-CoA affects this action.

Statins are currently the primary therapeutic modality for the management of hyperlipidemia. They inhibit HMG-CoA reductase (encoded by *HMGCR*), and reduce LDL-C levels by decreasing the intracellular cholesterol synthesis via the mevalonate pathway. As response to the lower intracellular cholesterol level, the gene expression of LDL-R is activated. However, the same transcription factor activating *LDLR* also regulates *PCSK9* (see Figure 1A). As result, correcting for this interaction between PCSK9 and HMGCR is essential for understanding the direct impact of PCSK9 on the lipid metabolism or any other complex outcome.

There has been mixed evidence for statins and lower cholesterol levels decreasing breast cancer (BC) risk. A meta-analysis of 2.4 million participants and 76,759 cases found no significant protective effect of statins against BC [6]. A clinical trial analysing disease free survival in pre-menopausal, stage I and II BC patients found improved survival in patients taking additionally zoledronic acid [7], which like statins inhibit the mevalonate pathway. Trials testing statins for BC survival are still underway [8] (see Figure 1B). Mendelian Randomization (MR) studies tested the effects of statins on BC risk by instrumenting variants at *HMGCR* associated with LDL-C [9,10], and found associations between genetically proxied inhibition of HMG-CoA reductase and reduced BC risk. However, there were no significant associations between variants at *PCSK9* or other lipid-lowering treatment targets and BC risk, or between genetically predicted LDL-C and BC risk.

PCSK9 does not only bind to LDL-R, but also to LDL-R related protein 1 (LRP1), very low-density lipoprotein receptor, apolipoprotein E receptor, and CD36 [11]. It has been shown that LDL-R effectively competes with LRP1 for PCSK9 activity [12], and lack of host PCSK9 reduced metastasis in liver in mice [13]. In a recent study, Mei et al. [14] identified a missense mutation, rs562556, in the *PCSK9* gene to be associated with BC metastasis and survival using four independent but small studies (combined sample size = 1,456). The proposed mechanism was by host PCSK9 binding to tumoral LRP1, which was suggested to affect metastatic colonization by inhibiting pro-metastatic gene expression. PCSK9 inhibitors reduced lung metastatic colonization in mice, and in human cell lines for lung and bone metastases [14]. It was suggested that the PCSK9 mechanism was not mediated by LDL-C reduction, and targeting homozygous carriers of the major allele might allow for early intervention strategies to prevent metastasizing BC in patients (see Figure 1B).

In this study, we explore this link between PCSK9 and BC survival further, with the main aim to replicate three findings of Mei et al. [14] (see also Figure 2): first, that rs562556 is associated with BC survival; second, that PCSK9 inhibition is beneficial for BC survival; and third, that the PCSK9 effect is independent of the cholesterol effect on BC survival. To achieve this, we use summary statistics from publicly available genome-wide association studies (GWASs), the UK biobank (UKB), and the FinnGen study. To analyze the causal link between PCSK9 and BC survival, we apply MR approaches, both univariable and multivariable, correcting for LDL-C levels.

**Figure 2:**
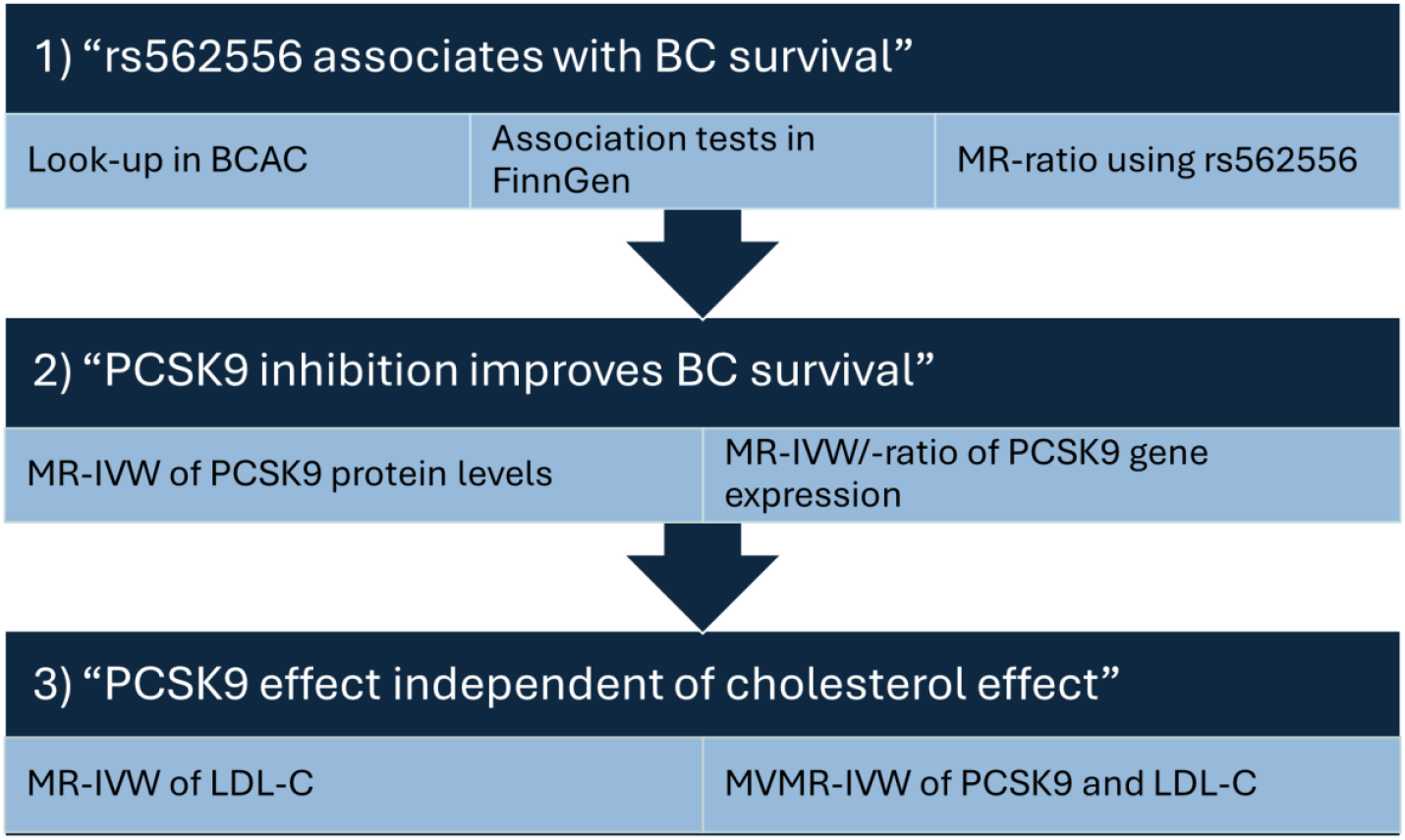
Overview of the three attempted replication analyses. The three statements are rephrased and simplified subsections from Mei et al. [14], and for each statement we list the methods used for the replication approach.

## Material and Methods

### Data sources

The analysis plan with data sources is displayed in Supplemental Figure S1. Publicly available genetic association summary statistics were used for most analyses. Appropriate ethical approvals and participant consent were obtained from the original studies that generated the data.

#### Exposure data

The primary exposure was PCSK9 protein levels in blood, as reported in our previous work of a sex-stratified meta-GWAS in Europeans [15]. As breast cancer survival is female-specific, we also focused on PCSK9 in females (n=8,936), and used the sex-combined data as sensitivity check (n= 20,016). The data of males and females was combined using the R package *meta* [16].

As a secondary exposure, we also obtained significant genetic association estimates on PCSK9 gene expression levels from GTEx v10 [17], which were available in 11 tissues. We excluded the tissue testis, as this male-specific tissue is not relevant for the female-specific outcomes considered here. To test and correct for LDL-C levels affecting BC survival, we downloaded both the female-specific and sex-combined GWAS summary statistics from the Global Lipids Genetics Consortium (GLGC, Europeans only, max. sample size females = 558,500; sex-combined = 1,231,284), as published in Kanoni et al. [18] and Graham et al. [19], respectively.

#### Primary outcome data

The primary outcome was BC survival, and data for this outcome was available in three studies. First, we used GWAS summary statistics from the Breast Cancer Association Cohort (BCAC), as published in Morra et al. [20]. They analyzed the survival of 91,686 breast cancer patients of European ancestry, with 7,531 breast cancer-specific deaths. Patients were over 18 at diagnosis with any stage of BC. In their GWAS, they used Cox regression to estimate the 15-year hazard ratios in an additive allele effect model.

Second, we used the SNP rs562556 summary statistics from Mei et al. [14] as provided in their supplemental Figure S1H (hazard ratios and p-values of rs562556 association with BC survival in four cohorts limited to European ancestry). No sample size information or case numbers were reported there. In two cohorts, patients were restricted to females over 50 at diagnosis with stage II or III breast cancer, hence enriched in patients with high risk of metastasis. Survival was censored at 10 years. In the other two studies, patients had metastatic BC and survival time was censored at 2 years. In their SNP tests per study, they used Cox proportional hazard regression with a recessive allele effect model. Here, we meta-analysed the four reported Hazard Ratios (HR) using the R package *meta* [16].

Given the differences in sample selection and tested SNPs, we obtained data from the FinnGen study, using the selection criteria from Mei et al., and testing all possible instruments for association. The FinnGen study is a large-scale genomics initiative that has analysed over 500,000 Finnish biobank samples and correlated genetic variation with health data to understand disease mechanisms and predispositions. The project is a collaboration between research organisations and biobanks within Finland and international industry partners [21]. The FinnGen project possesses the ethical and prior permits required for biobank research (see Supplementary Note). All participants have provided written consent to biobank research, either in connection with sample donation or when taking part in previous research projects whose materials have been transferred to Finnish biobanks with the approval of Fimea, the National Supervisory Authority for Welfare and Health. Information on genotyping and imputation can be found in the Supplementary Note. The FinnGen data freeze R12 was utilised, and female breast cancer patients were identified based on The International Classification of Diseases, Tenth Revision (ICD-10) code C50. Only patients who had stage II or stage III breast cancer and who were older than 50 at the time of diagnosis were included in the analysis. The final study sample consisted of 4648 breast cancer patients.

#### Secondary outcome data

We repeated the MR analyses on positive and negative controls. As negative control, we used BC incidence, as current literature does not link PCSK9 or LDL-C reduction by PCSK9 inhibition to BC risk. Data was taken from the FinnGen and UK Biobank (UKB) meta-analysis (cases = 30,593, controls = 388,840) [21], and we expected to find a null-result here. As positive control we used parental age at death (PAD) as a proxy for own survival (data source: UKB, n= 208,118) [22], and coronary artery disease (CAD, data source: Aragam et al.; meta-analysis of CARDIoGRAMplusC4D, UKB, nine other studies) [23]. CAD summary statistics were available both sex-combined and females only (cases = 181,522, controls = 984,168 in the sex-combined setting). PCSK9 and LDL-C affect survival by increasing the risk for cardiovascular events. Hence, increasing levels of PCSK9 or LDL-C are expected to have a significant positive effect on CAD, and a significant, negative effect on survival, resulting in decreased parental age at death.

### Instrumental variables

For the pQTLs, we used the same for SNPs as described in Pott et al. [15], namely rs11591147, rs693668, rs11583680, and rs2495491. They all reached genome-wide significance (*p*<5×10^−8^) and are pairwise independent (LD *r*^2^<0.1).

For the gene expression, we filtered the GTEx v10 data set for genome-wide significant associations or significant association for rs562556 (nominal p-value threshold per tissue taken from GTEx, max p-value threshold 1.9×10^−4^), and found initially 12 tissues with significant eQTLs. We excluded testis and eQTLs with no matching outcome data. This left us with 9 tissues for MR analyses (adipose visceral omentum, brain cerebellar hemisphere, brain cerebellum, Esophagus muscularis, lung, nerve tibial, pancreas, spleen, and whole blood). For each tissue, we selected independent variants after clumping to a pairwise LD threshold of *r*^2^<0.1 using the 1000 Genome European reference panel assessed with FUMA [24].

For LDL-C, we restricted the GLGC data to genome-wide significant SNPs after genomic control correction, minor allele frequency (MAF) >0.01, and annotated with rsID. We further reduced to variants also available in the outcome data sets. We then used position based priority pruning by selecting the best-associated (highest absolute Z-score) variant as index SNP, and excluding all variants within +/-500 MB as tagged SNPs. Finally, we only used index SNPs with strong support by excluding those with less than 9 tagged SNPs. This resulted in 171 and 269 independent index SNPs for the MR analyses for females and sex-combined setting, respectively. In addition to the genetically proxied LDL-C levels, we also selected instruments to proxy HMG-CoA reductase or PCSK9 inhibition. For this we used independent SNPs at *HMGCR* and *PCSK9* as reported by Yang et al. (eTable 3) [25].

We harmonized the effect allele to be the minor allele throughout, and excluded tri-allelic SNPs and SNPs with effect allele frequency difference >0.1 between exposure and outcome. A total of 405 SNPs were selected, pairwise independent for their respective exposure.

### FinnGen genotyping and statistical analysis

Cox regression with an additive model was utilised to estimate hazard ratios and 95% confidence intervals for 403 predefined SNPs, with age used as a covariate. In addition, we performed a Cox regression with a recessive model for variant rs562556 using age as a covariate, as in our main analysis. The time-to-event was censored at the time of last follow-up or at 10 years after diagnosis, whichever came first. Patients who died from causes other than breast cancer were censored at the time of death if death occurred before 10 years from diagnosis or at 10 years if they did not. A total of 288 breast cancer deaths occurred out of 4648 patients with the disease.

For five SNPs, the allele frequency was very low in the FinnGen study (AF<0.01). Hence we excluded those five variants. All 398 used instruments and their summary statistics on exposures and outcomes be found in the Supplemental Tables S1 (exposure association for the MR approach), S2 (exposure association for the MVMR approach), and S3 (outcome association for both MR and MVMR approaches).

### Mendelian randomization

We tested for causal effects of PCSK9 and LDL-C levels on BC survival and all secondary outcomes (BC risk, parental longevity, and CAD) using three MR approaches. First, we estimated the Wald ratio and standard error from the first two terms of the delta method [26] using the SNP rs562556, as it was suggested as causal variant for BC survival. We choose the first two terms of the delta method to address potential violation of the no measurement error assumption due to weak instruments, as the p-value threshold was here relaxed to *p*<0.05. Second, we combined the available instrument per subgroup or tissue using inverse variance weighted MR (MR-IVW). The method was used as implemented in the R package *MendelianRandomization* [27]. When only one instrument was available, the simple ratio and standard error from the first two terms of the delta method were estimated [26]. For LDL-C, we ran the MR-IVW using either any available instrument, SNPs at *PCSK9* to mimic PCSK9 inhibition, SNPs at *HMGCR* to mimic statin treatment, or SNPs at *PCSK9* and *HMGCR*. We corrected for multiple testing using the Benjamini & Hochberg procedure [28].

In a third approach, we estimate the PCSK9 effect conditional to the LDL-C effect using multivariable MR (MVMR). The MVMR-IVW was performed using the same R-package, and we used the maximum sample size per exposure to estimate the conditional F-statistics. We tested the MVMR using either all four instruments from the pQTL MR approach or SNPS at *PCSK9* and *HMGCR* from the LDL-C approach. Other combination such as all valid LDL-C variants would result in weak instruments for PCSK9. We applied the same false discovery correction as for the MR approaches.

## Results

### Replication 1: missense mutation rs562556 on BC survival

Mei et al. reported a significant association between the PCSK9 missense mutation rs562556 and BC survival. We looked this SNP up in the publicly available BCAC data set and additionally analysed it in FinnGen. The results are summarised in Table 1. The association did not reach significance in neither BCAC nor FinnGen. This was irrespective of the genetic model used to analyse the effect.

**Table 1:**
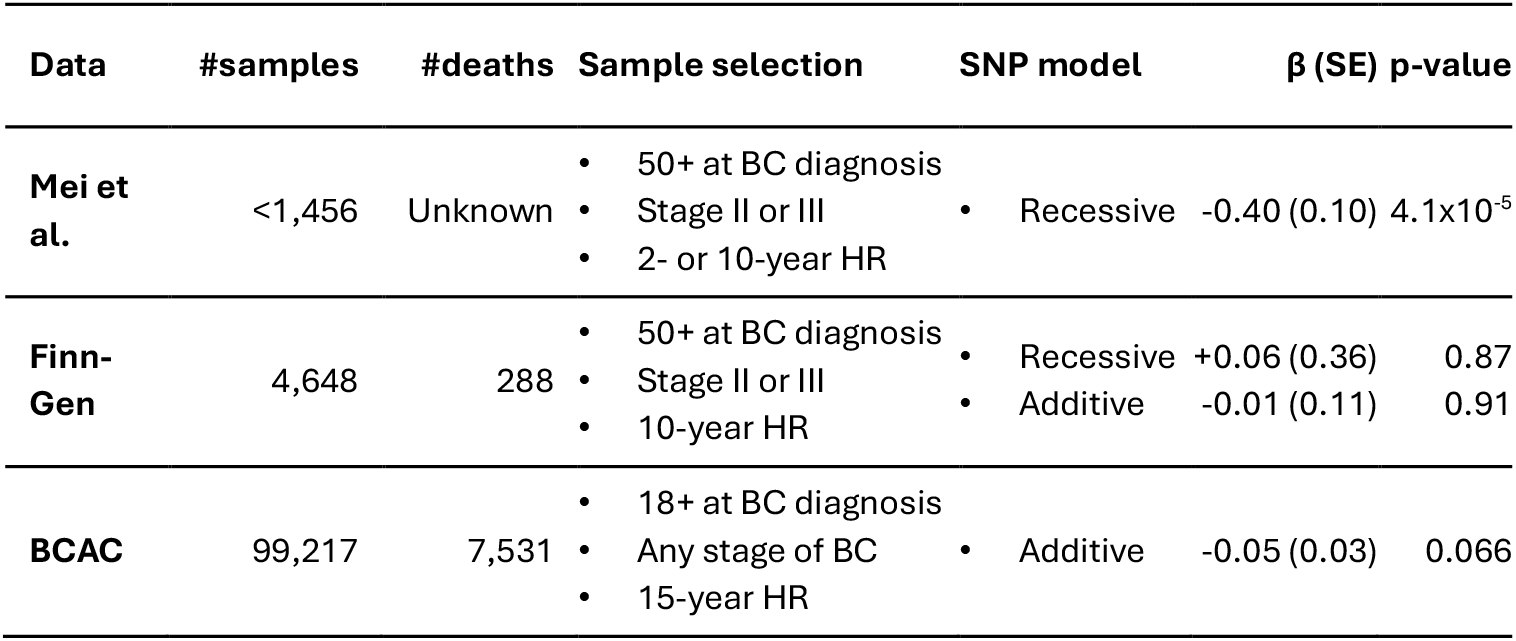
Overview of used outcome data sources for BC Survival. Mei et al. *[14]* used four small BC cohorts testing common human germline missense variants only. The PCSK9 variant rs562556 was detected in the recessive model only. The sample selection of Mei et al. was used to select BC patients in the FinnGen to allow an replication approach most similar to Mei et al. The BC Association Consortium (BCAC) used a genome-wide approach and different sample selection.

We used rs562556 in the MR-ratio approach to test for significant association between PCSK9 levels and BC survival, using all four outcome statistics. Significant exposure associations were available for PCSK9 protein levels (females only and sex-combined) and *PCSK9* gene expression in spleen and esophagus (muscularis) tissue, with F-statistics >10 (16.3, 63.7, 20, and 18.9, respectively). We observed significant positive association between PCSK9 levels and BC survival when using the Mei et al. outcome data (e.g. PCSK9 in females: *logHR* = 18.03, *p* = 3.99×10^−3^), but not when using data from BCAC or FinnGen (see Figure 3A, Supplemental Figure S2 and Supplemental Table S4). We note that these analyses using Mei et al data are not independent, nor are these true replications of the previously published finding, as they are based on the same outcome associations.

**Figure 3:**
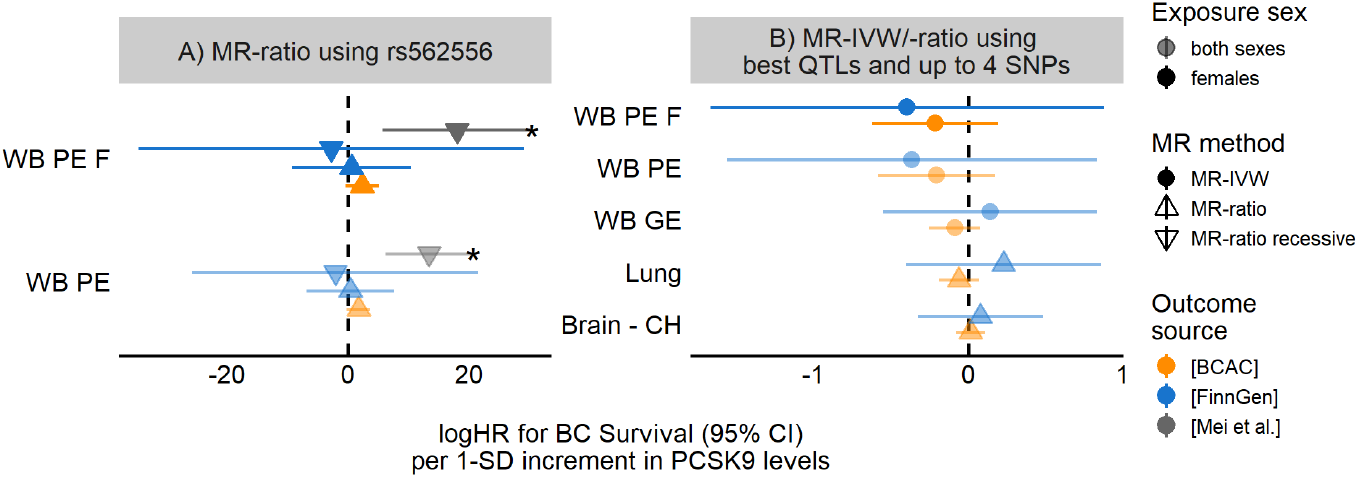
Forest plot of the MR analyses on BC survival. The log hazard ratios (logHR) for BC survival per 1 SD increment in PCSK9 levels are given for protein expression (PE) and selected tissues (gene expression, GE). A) MR-ratio results using rs562556, representing the functionality of PCSK9 to bind to LRP1. B) MR-IVW or MR-ratio results are shown, using the best QTLs per subgroup or tissue. The IVW and ratio estimates were not significantly different from 0 in when testing PCSK9 on the BCAC or FinnGen data, but significant when using data from the small studies reported by Mei et al. *[14]*. * significant after multiple testing correction; WB – whole blood; PE – protein expression; F – females only; CH – cerebellar hemisphere

### Replication 2: PCSK9 levels on BC survival

Mei et al. [14] also reported a significant reduction of metastasis due to PCSK9 inhibition. To replicate these findings, we used multiple independent variants at the *PCSK9* gene region associated with PCSK9 protein levels or gene expression. Here, we only used outcome data from FinnGen and BCAC. The analysis was well powered again, with F-statistics ranging from 32.1 (gene expression in pancreatic tissue) to 310.6 (protein levels sex-combined). The results are summarized in Figure 3B, Supplemental Figure S3 and Supplemental Table S5. Again, we did not observe significant association between genetically predicted PCSK9 levels and BC survival (e.g. PCSK9 in females: *logHR* = −0.21 in BCAC; *logHR* = −0.39 in FinnGen, both *p* > 0.05, see also scatter plot in Supplemental Figure S4A). Similarly, the effect of genetically predicted PCSK9 gene expression on BC survival was not significant and with mixed effect directions, depending on tissue and outcome source.

### Replication 3: Impact of LDL-C on PCSK9 effect

Mei et al. [14] suggested that the PCSK9 effect would be independent from the effect of the lipid metabolism. To test this hypothesis, we first analysed the effect of LDL-C on BC survival with the MR-IVW approach, and then used the MVMR-IVW approach to estimate the effect of PCSK9 on BC survival, conditional on the LDL-C effect. In Table 2, we provide a comparison of the results for LDL-C and PCSK9 per MR-approach and SNP selection.

**Table 2:**
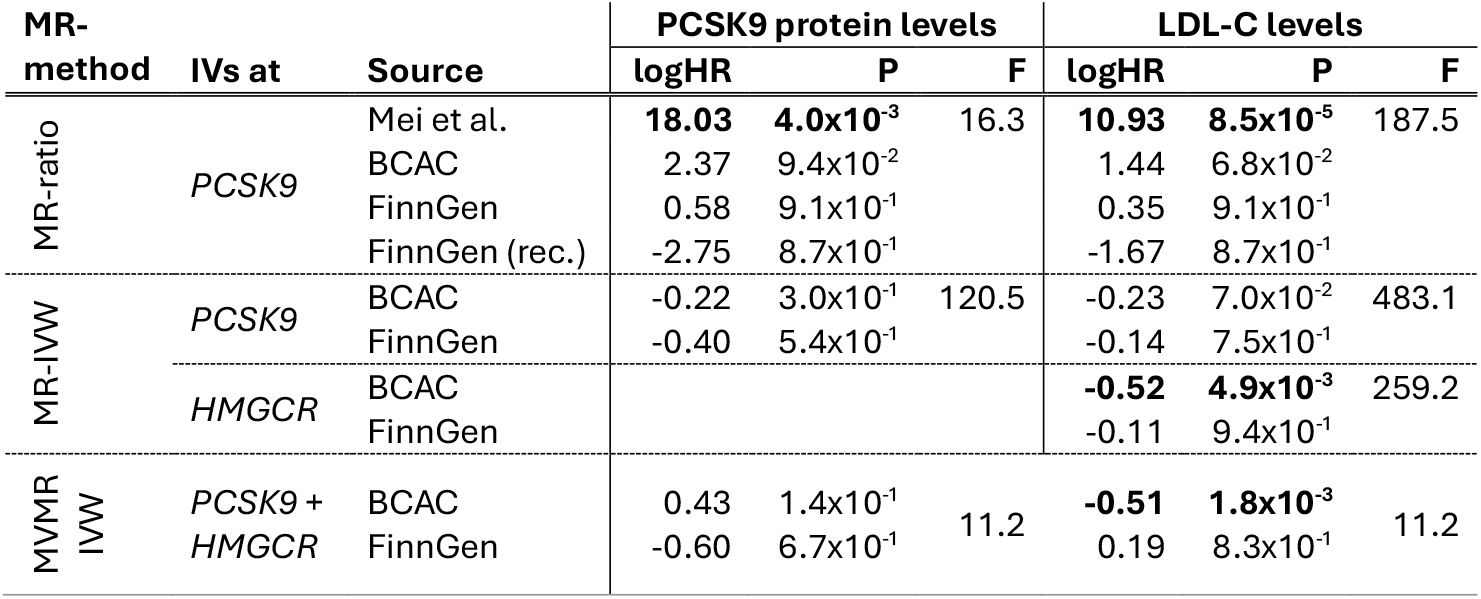
Overview of the MR results of PCSK9 and LDL-C levels on BC survival in females. In the MR-ratio method, only rs562556 at PCSK9 was used. In the MR-IVW method, we report here the results using instruments at *PCSK9* and/or *HMGCR*. The latter was only available for LDL-C, as no valid instruments for PCSK9 were observed there. In the MVMR, we combined the instruments at *PCSK9* and *HMGCR*. logHR: log Hazard Ratio; P: p-value; F: (conditional) F-statistic; rec: recessive genetic model.

For the LDL-C MR-IVW approaches, the F-statistics ranged from 259.2 (instruments only at *HMGCR*) to 622.3 (instruments only at *PCSK9*). When using all available instruments in the MR-IVW, we observed no significant result (e.g. LDL-C in females: *logHR* = 0.005, *p* = 0.877 for BCAC). As expected, when using 12 instruments from the *PCSK9* gene region, the results for LDL-C were quite similar to the ones for PCSK9 protein levels: *logHR* = −0.23 (*p* = 0.070) for LDL-C in females for BCAC. Although the analyses using LDL-C as exposure were better powered, the estimates did not reach significance. Finally, when using instruments at the *HMGCR* gene region, we observed a significant negative association with BC survival in BCAC (e.g. LDL-C in females: *logHR* = −0.514, *p* = 4.38×10^−3^, see also Supplemental Figure S5A and S5B), but not in the FinnGen data (*logHR* = −0.107, *p* = 0.94). In other words, genetically proxied inhibition of HMG-CoA reductase (e.g. statin treatment) has mixed results regarding survival probability in breast cancer patients. A summary of the results is shown in Figure 4A, and all results are given in Supplemental Table S5.

**Figure 4:**
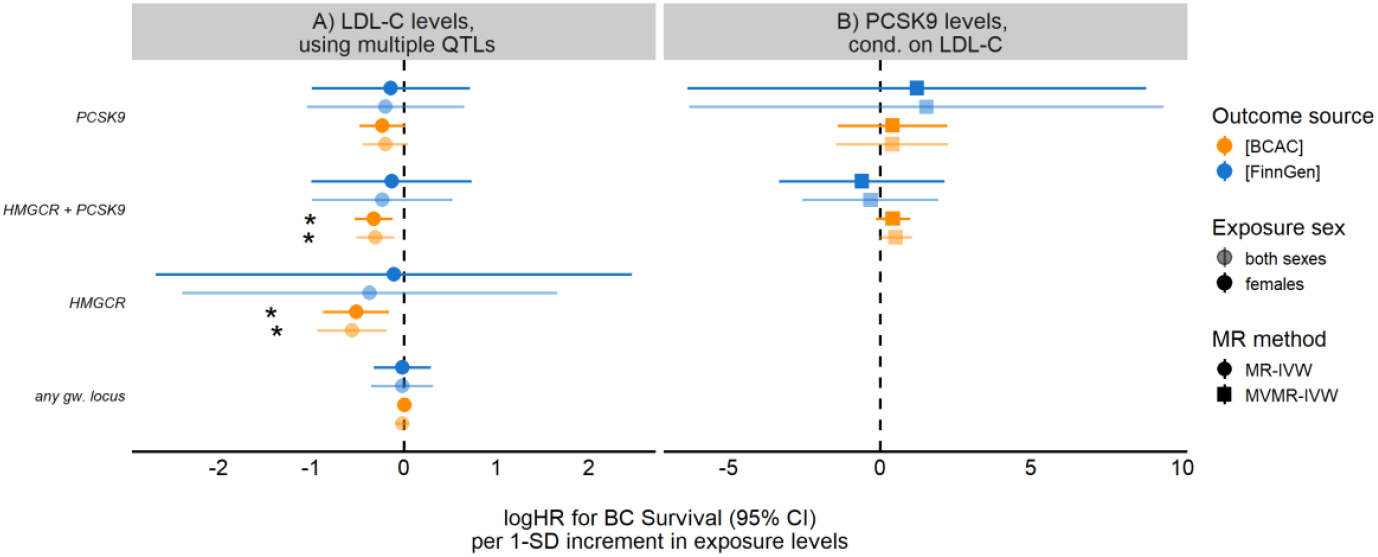
Forest plots of the MR analyses of LDL-C and of the MVMR analyses of PCSK9 conditional on LDL-C on BC survival. The log hazard ratios (logHR) for breast cancer survival per 1 SD increment in PCSK9 or LDL-C levels are given per selected instruments. A) The LDL-C effect was negative in the univariable MR-IVW, and reached significance when including *HMGCR* instruments and the BCAC data set. B) The MVMR-IVW PCSK9 effect was estimated as positive in BCAC, but negative in FinnGen when using both *HMGCR* and *PCSK9* instruments. However, PCSK9 effects do not reach significance. * significant after multiple testing correction.

To correct for potential influence of LDL-C on the PCSK9 effect on BC survival, we then tested both PCSK9 and LDL-C on BC survival with MVMR-IVW. Using the SNPs at *PCSK9* and *HMGCR* and BCAC outcome data, we found that the effect of LDL-C on BC survival stayed similar to the univariable estimate for *HMGCR* (*logHR* = −0.51, *p* = 1.8×10^−3^), while the effect of PCSK9 changed its direction but remained non-significant (females: *logHR* = 0.30, *p* = 0.29, conditional F-statistic = 11.1; sex-combined: *logHR* = 0.28, *p* = 0.06 conditional F-statistic = 15.2). However, using the FinnGen data, the LDL-C effect changed direction, while the PCSK9 effect stayed negative and non-significant. Using instruments at *PCSK9* only led to a reduction of power, with conditional F-statistic of PCSK9 of 9.2. As result, all the MVMR estimates were not significant. Results are shown in Table 2, Figure 4B and in Supplemental Table S6.

### Sensitivity checks with BC risk, longevity, and CAD

We repeated our analyses for three other outcomes: BC risk, parental age at death as proxy for overall survival, and risk for CAD. Forest plots are given in Figure S6 and S7, and all results are summarized in Supplemental Tables S4 – S6 for the MR-ratio of rs562556, MR-IVW/MR-ratio of QTLs, and MVMR-IVW, respectively.

BC risk acted as negative control, and indeed none of the breast cancer tests were significant, e.g. no causal effect of PCSK9 protein or PCSK9 gene expression levels on risk of BC (see Supplemental Figure S4B and S6). For genetically proxied LDL-C levels there was no effect (*logOR* = 0.01, *p* = 0.586 in females), but genetically proxied HMG-CoA reductase activity was significantly associated with BC risk (*logOR* = 0.20, *p* = 0.039). However, after multiple testing correction, this association was no longer significant (*p*_*adj*_= 0.108). In the MVMR approach, the results were not significant. However, the effect of LDL-C was still estimated positively (*logOR* = 0.171, *p* = 0.053 using sex-combined exposure data and instruments at both *PCSK9* and *HMGCR*), while the PCSK9 effect was estimated as negative (*logOR* = −0.239, *p* = 0.068).

CAD risk and PAD were used as positive control, and as expected almost all MR analyses with CAD (sex-combined) were significant with a positive effect direction (see Supplemental Figure S4D and S6). Only the effect of PCSK9 gene expression in pancreas tissue did not reach significance (*p* = 0.056). The effects on CAD in females were also positive, but due to lower sample size and case number not always significant after correction. Of note, the association between *HMGCR* SNPs and CAD in females was markedly reduced. For example, rs12916, a well-known variant in the 3’ UTR of *HMGCR* is strongly associated with LDL-C in both females and sex-combined and with CAD in the sex-combined setting, but not in the females only setting (*logOR* = 0.028, *p* = 1×10^−8^ sex-combined, *logOR* = 0.006, *p* = 0.615 in females, see also Supplemental Table S3). Hence, the results of LDL-C proxied by HMG-CoA reductase activity on CAD was not significant in females (*logOR* = 0.097, *p* = 0.521 in females vs. *logOR* = 0.382, *p* = 1×10^−6^ sex-combined, see also Supplemental Figure S5C and S5D). Another consequence of the CAD differences was found in the MVMR: we observed significant effects of PCSK9 on CAD conditional on LDL-C when using CAD sex-combined outcome data, but not in the CAD females only data (see Supplemental Figure S7). For PCSK9, the effect sizes were similar, but the uncertainty larger in females (MVMR *logOR* = 0.290, *p* = 0.136 for PCSK9 on CAD females, *logOR* = 0.269, *p* = 3.49×10^−3^ for PCSK9 on CAD sex-combined). For the LDL-C effect, the sizes also differed (MVMR *logOR* = 0.252, *p* = 3.83×10^−2^ for LDL-C on CAD females, *logOR* = 0.436, *p* = 4.44×10^−14^ for LDL-C on CAD sex-combined). The effects on PAD were also significant for PCSK9 protein and LDL-C levels (see Supplemental Figure S4C), but failed at all gene expression tissues and for the *HMGCR* instruments. No significant results were found in the MVMR approach.

## Discussion

In this study, we tried to replicate the findings of Mei et al. [14], who observed a significant effect of the PCSK9 loss-of-function variant rs562556 on breast cancer metastatic colonization, and suggested that PCSK9 inhibition might improve survival chances in breast cancer patients homozygous for the major allele of the missense mutation rs562556. We used Mendelian Randomization to estimate causal effects of life-time exposure of PCSK9, and tested for breast cancer survival, using data from the breast cancer association consortium (BCAC) [20].

PCSK9 inhibitors are administered subcutaneously, and then absorbed into the bloodstream. They are mainly distributed through the circulatory system, with minimal extravascular distribution. Hence, to test if PCSK9 inhibitors could affect breast cancer survival, we used genetically predicted PCSK9 protein levels from a large meta-GWAS [15]. Even with the strong power that comes from cis-effects, we observed no significant effect in the MR-IVW approach with negative point estimates. In the MVMR-IVW approach, we corrected for mediating effects through the lipid metabolism and estimated the direct effect of PCSK9 on BC survival. Here, the effect direction changed, but still failed to reach significance. Finally, we used the reported missense mutation rs562556 in a ratio estimation approach. Here, we used the effects of rs562556 as reported in the BCAC, in FinnGen and by Mei et al. [14], and we detected significant positive effects of PCSK9 on breast cancer survival only when using the data from Mei et al. When using rs562556 data from the BCAC or FinnGen, the effect was positive, but not significant. This was true for both the protein expression subgroups and the tissue-wise gene expression. The differences in effect size and significance could be explained by several things, which we will discuss in the following.

For the IVW approaches, we used instruments with strong effects on PCSK9 levels. Their direct effect on PCSK9 levels has been previously described by others [15], and can be explained by their role in gene expression, splicing regulation, and increased protein degradation. In contrast, the missense mutation rs562556 has not been described as driver for PCSK9 levels but its functionality. In line with this, Mei et al. described V474I, the mutation caused by rs562556, to have a higher binding affinity to tumoral LRP1, which in turn activates pro-metastatic gene expression and increases the risk for metastasis. However, in their mouse and cell line models they could also show a benefit by blocking any PCSK9 by monoclonal antibodies. Hence the two MR approaches with their separate instruments might represent different pathways of PCSK9 action, either by levels or function. The different scaling and significance in the MR-ratio estimates could be explained by several things, which we discuss in the following.

First, Morra et al. [20] and Mei et al. [14] used different genetic models to estimate the effect of rs562556 on breast cancer survival: additive allele effect or recessive effect model, respectively. The choice of genetic model can affect the observed SNP effect estimate size and hence the ratio estimate size. In FinnGen, both genetic models were tested, and in both tests the effect of rs562556 was not significant on BC survival. In the recessive model, which represents the replication of the SNP effect, we also observed a different effect direction than reported by Mei et al. [14].

Second, all three studies used the cox regression model to estimate the average HR of the exposed versus unexposed. For rs562556, this mean higher vs lower binding affinity of PCSK9 to LRP1, while for the PCSK9 QTLs it stands for higher versus lower levels of PCSK9. However, the estimated effect size depends on the considered survival time, as the average HR ignores the distribution of events during the follow-up [29]. In our case, this time-variability of HR might have caused the difference in effect sizes, as Morra et al. [20] were looking at 15-year survival, while Mei et al. [14] considered 10-year survival for two cohorts with high risk patients and 2-year survival for two cohorts of patients with metastasis. In addition, Mei et al. [14] stratified their sample sets by including only patients with high risk for metastasis: females over 50 years of age at diagnosis with stage II or III breast cancer. This sample stratification can potentially induce bias in the HR estimates, and limit generalization to low risk patients as included in Morra et al. [20], e.g. females over 18 with any stage of BC. We repeated all analyses in FinnGen, which used the same sample selection as described in Mei et al. [14]. However, even then we could not detect a significant association of PCSK9 levels on BC survival.

Third, the effect could be modified by LDL-C. Mei et al. reported an effect independent of the lipid metabolism, and other studies suggest that statins could be beneficial during breast cancer treatment [8]. We observed significant negative effects logHR for LDL-C on BC survival when using instruments at *HMGCR*. Statins, which inhibit the HMG-CoA activity and lower LDL-C, are known to also increase PCSK9 levels. Hence, there might be an unobserved interaction between *PCSK9, HMGCR* and LDL-C on BC survival. We attempted to unravel this interaction by applying multivariable MR. The effect direction changed, but remained unsignificant. However, the effects at *PCSK9* are highly correlated, and the power to detect effects in the MVMR was limited: the conditional F-statistics were 11.2, while in the MR approach we had F-statistics of 120.5 and 393.5 for PCSK9 and LDL-C, respectively.

Finally, the association in Mei et al. [14] may have been a false positive finding. Although the SNP effect validated in mouse knock-out studies, it was not replicated in the larger dataset of the BCAC or FinnGen. It is possible that PCSK9 inhibitors might have a beneficial effect in mouse models, cell line, or high-risk BC patients, but the generalization to all patient groups might not be valid.

To ensure our instruments and exposures were well defined, we ran positive and negative controls. For BC risk, we did not expect to find any effect of PCSK9 or LDL-C using instruments at *PCSK9* or genome-wide [6,10], and indeed we found none. However, we were able to replicate the known effect of genetically proxied HMG-CoA reductase inhibition (e.g. statin treatment) [9,10]. The positive control was CAD risk and parental age at death, and the PCSK9 effects were according to expectation positive on CAD and negative on parental age at death.

One limitation of this study was that we could not exactly test Mei et al.’s suggestion that PCSK9 inhibitors would be beneficial for homozygous major allele carriers, which would focus more on PCSK9 functionality rather than just PCSK9 levels. However, the SNPs used in the MR-IVW approach were rather independent of rs562556, with LD r^2^<0.05 for all but two variants: rs553741 (r^2^=0.374, eQTL in liver) and rs693668 (r^2^=0.235, pQTL). Given this independence, it is rather unlikely that the effect directions will change when restricting the GWAS on the subset of samples carrying the major allele. The SNP rs693668 has been reported for sex-biased effects, with stronger effects on PCSK9 gene expression in women [30], but stronger effects on PCSK9 protein expression in men [15]. The mechanism is not yet clear, although it has been hypothesized that estradiol might regulate PCSK9 expression [31]. Hence, it would be of future interest to test the MR with outcome data in breast cancer patients stratified for estrogen receptor status (ER+ vs ER-). Another limitation is that MR tests for life-long effects, and might be of limited usage for understanding drug effects. Also, we cannot exclude potential drug interactions between chemotherapy and statin treatment, which might explain the beneficial effects observed in animal models and cell lines.

In conclusion, a significant positive effect of PCSK9 on breast cancer survival could only be reproduced when using the exact same outcome data set from Mei et al. [14], but not when using independent data from a consortium [20] or the FinnGen [21].

## Data Availability

All data produced in the present work are contained in the manuscript

https://zenodo.org/records/10600167

https://console.cloud.google.com/storage/browser/gtex-resources

https://csg.sph.umich.edu/willer/public/glgc-lipids2021/results/sex_and_ancestry_specific_summary_stats/

https://www.dropbox.com/scl/fi/i4lncx5l6r3pxy5c4ghig/bcac_survival_results_2021.csv.gz?rlkey=p30butd2aqengfp0g16ljwmso&st=rrcl27at&dl=0

https://console.cloud.google.com/storage/browser/finngen-public-data-r10/ukbb/

https://hugeamp.org/dinspector.html?dataset=Aragam2022_CAD_Mixed_females&phenotype=CAD

https://hugeamp.org/dinspector.html?dataset=Aragam2022_CAD_EU&phenotype=CAD

https://www.ebi.ac.uk/gwas/studies/GCST006702

## Declarations

### Availability of data and code

The code is available on https://github.com/pottj/PCSK9_BreastCancerSurvival. We used R v4.2.2 and the R-package MendelianRandomization v0.10.0 [27] throughout the analyses. All extracted and used summary statistics are available in the Supplemental Table S1 – S3.

- GWAS summary statistics on PCSK9 plasma levels has been contributed by Pott et al. [15]. and has been downloaded from https://zenodo.org/records/10600167.
- Genetic summary statistics on PCSK9 gene expression has been contributed by GTEx consortium [17] and has been downloaded from https://console.cloud.google.com/storage/browser/gtex-resources.
- GWAS summary statistics on LDL-C levels has been contributed by the Global Lipids Genetics Consortium [18,19] and has been downloaded from https://csg.sph.umich.edu/willer/public/glgc-lipids2021/results/sex_and_ancestry_specific_summary_stats/.
- GWAS summary statistics on breast cancer survival has been contributed by the Breast Cancer Association Consortium [20] and has been downloaded from https://www.dropbox.com/scl/fi/i4lncx5l6r3pxy5c4ghig/bcac_survival_results_2021.csv.gz?rlkey=p30butd2aqengfp0g16ljwmso&st=rrcl27at&dl=0.
- GWAS summary statistics on breast cancer has been contributed by FinnGen and UK Biobank [21] and has been downloaded from https://console.cloud.google.com/storage/browser/finngen-public-data-r10/ukbb/.
- GWAS summary statistics on coronary artery disease has been contributed by Aragam et al. [23], and has been downloaded from https://hugeamp.org/dinspector.html?dataset=Aragam2022_CAD_Mixed_females&phenotype=CAD and https://hugeamp.org/dinspector.html?dataset=Aragam2022_CAD_EU&phenotype=CAD
- GWAS summary statistics on parental age at death has been contributed by Pilling et al. [22], and has been downloaded from https://www.ebi.ac.uk/gwas/studies/GCST006702
- In FinnGen [21], the individual-level data are available under restricted access for legal and ethical reasons. Formal approval for the researchers is required to access the data: please see https://www.finngen.fi/en/access_results for more details. To get access to FinnGen summary statistics, fill out an online form at: https://elomake.helsinki.fi/lomakkeet/124935/lomake.html. Access to individual-level data and genotype data is managed by the Finnish Biobank Cooperative at the Fingenious portal [https://site.fingenious.fi/en/]). The expected response time for access requests to individual-level data is 1-2 months.

## Acknowledgements

We want to acknowledge the participants and investigators of FinnGen study.

## References

1. Norata GD, Tavori H, Pirillo A, Fazio S, Catapano AL. Biology of proprotein convertase subtilisin kexin 9: beyond low-density lipoprotein cholesterol lowering. Cardiovasc Res. 2016;112: 429–442. doi:10.1093/cvr/cvw194

2. Zhang D-W, Garuti R, Tang W-J, Cohen JC, Hobbs HH. Structural requirements for PCSK9-mediated degradation of the low-density lipoprotein receptor. Proc Natl Acad Sci U S A. 2008;105: 13045–13050. doi:10.1073/pnas.0806312105

3. Sabatine MS, Giugliano RP, Wiviott SD, Raal FJ, Blom DJ, Robinson J, et al. Efficacy and Safety of Evolocumab in Reducing Lipids and Cardiovascular Events. N Engl J Med. 2015;372: 1500–1509. doi:10.1056/NEJMoa1500858

4. Cannon CP, Cariou B, Blom D, McKenney JM, Lorenzato C, Pordy R, et al. Efficacy and safety of alirocumab in high cardiovascular risk patients with inadequately controlled hypercholesterolaemia on maximally tolerated doses of statins: the ODYSSEY COMBO II randomized controlled trial. Eur Heart J. 2015;36: 1186–1194. doi:10.1093/eurheartj/ehv028

5. Lamb YN. Inclisiran: First Approval. Drugs. 2021;81: 389–395. doi:10.1007/s40265-021-01473-6

6. Undela K, Srikanth V, Bansal D. Statin use and risk of breast cancer: a meta-analysis of observational studies. Breast Cancer Res Treat. 2012;135: 261–269. doi:10.1007/s10549-012-2154-x

7. Gnant M, Mlineritsch B, Stoeger H, Luschin-Ebengreuth G, Heck D, Menzel C, et al. Adjuvant endocrine therapy plus zoledronic acid in premenopausal women with early-stage breast cancer: 62-month follow-up from the ABCSG-12 randomised trial. Lancet Oncol. 2011;12: 631–641. doi:10.1016/S1470-2045(11)70122-X

8. Zipinotti Dos Santos D, De Souza JC, Pimenta TM, Da Silva Martins B, Junior RSR, Butzene SMS, et al. The impact of lipid metabolism on breast cancer: a review about its role in tumorigenesis and immune escape. Cell Commun Signal. 2023;21: 161. doi:10.1186/s12964-023-01178-1

9. Carter P, Vithayathil M, Kar S, Potluri R, Mason AM, Larsson SC, et al. Predicting the effect of statins on cancer risk using genetic variants from a Mendelian randomization study in the UK Biobank. Elife. 2020;9: e57191. doi:10.7554/eLife.57191

10. Sun L, Ding H, Jia Y, Shi M, Guo D, Yang P, et al. Associations of genetically proxied inhibition of HMG-CoA reductase, NPC1L1, and PCSK9 with breast cancer and prostate cancer. Breast Cancer Res. 2022;24. doi:10.1186/s13058-022-01508-0

11. Schulz R, Schlüter K-D. PCSK9 targets important for lipid metabolism. Clin Res Cardiol Suppl. 2017;12: 2–11. doi:10.1007/s11789-017-0085-0

12. Canuel M, Sun X, Asselin M-C, Paramithiotis E, Prat A, Seidah NG. Proprotein Convertase Subtilisin/Kexin Type 9 (PCSK9) Can Mediate Degradation of the Low Density Lipoprotein Receptor-Related Protein 1 (LRP-1). Kanzaki M, editor. PLoS ONE. 2013;8: e64145. doi:10.1371/journal.pone.0064145

13. Sun X, Essalmani R, Day R, Khatib AM, Seidah NG, Prat A. Proprotein Convertase Subtilisin/Kexin Type 9 Deficiency Reduces Melanoma Metastasis in Liver. Neoplasia. 2012;14: 1122–IN5. doi:10.1593/neo.121252

14. Mei W, Faraj Tabrizi S, Godina C, Lovisa AF, Isaksson K, Jernström H, et al. A commonly inherited human PCSK9 germline variant drives breast cancer metastasis via LRP1 receptor. Cell. 2025;188: 371-389.e28. doi:10.1016/j.cell.2024.11.009

15. Pott J, Kheirkhah A, Gadin JR, Kleber ME, Delgado GE, Kirsten H, et al. Sex and statin-related genetic associations at the PCSK9 gene locus: results of genome-wide association meta-analysis. Biol Sex Differ. 2024;15: 26. doi:10.1186/s13293-024-00602-6

16. Balduzzi S, Rücker G, Schwarzer G. How to perform a meta-analysis with R: a practical tutorial. Evid Based Mental Health. 2019;22: 153–160. doi:10.1136/ebmental-2019-300117

17. The GTEx Consortium, Aguet F, Anand S, Ardlie KG, Gabriel S, Getz GA, et al. The GTEx Consortium atlas of genetic regulatory effects across human tissues. Science. 2020;369: 1318– 1330. doi:10.1126/science.aaz1776

18. Kanoni S, Graham SE, Wang Y, Surakka I, Ramdas S, Zhu X, et al. Implicating genes, pleiotropy, and sexual dimorphism at blood lipid loci through multi-ancestry meta-analysis. Genome Biol. 2022;23: 268. doi:10.1186/s13059-022-02837-1

19. Graham SE, Clarke SL, Wu K-HH, Kanoni S, Zajac GJM, Ramdas S, et al. The power of genetic diversity in genome-wide association studies of lipids. Nature. 2021;600: 675–679. doi:10.1038/s41586-021-04064-3

20. Morra A, Escala-Garcia M, Beesley J, Keeman R, Canisius S, Ahearn TU, et al. Association of germline genetic variants with breast cancer-specific survival in patient subgroups defined by clinic-pathological variables related to tumor biology and type of systemic treatment. Breast Cancer Res. 2021;23: 86. doi:10.1186/s13058-021-01450-7

21. Kurki MI, Karjalainen J, Palta P, Sipilä TP, Kristiansson K, Donner KM, et al. FinnGen provides genetic insights from a well-phenotyped isolated population. Nature. 2023;613: 508– 518. doi:10.1038/s41586-022-05473-8

22. Pilling LC, Kuo C-L, Sicinski K, Tamosauskaite J, Kuchel GA, Harries LW, et al. Human longevity: 25 genetic loci associated in 389,166 UK biobank participants. Aging. 2017;9: 2504– 2520. doi:10.18632/aging.101334

23. Aragam KG, Jiang T, Goel A, Kanoni S, Wolford BN, Atri DS, et al. Discovery and systematic characterization of risk variants and genes for coronary artery disease in over a million participants. Nat Genet. 2022;54: 1803–1815. doi:10.1038/s41588-022-01233-6

24. Watanabe K, Taskesen E, Van Bochoven A, Posthuma D. Functional mapping and annotation of genetic associations with FUMA. Nat Commun. 2017;8: 1826. doi:10.1038/s41467-017-01261-5

25. Yang G, Mason AM, Gill D, Schooling CM, Burgess S. Multi-biobank Mendelian randomization analyses identify opposing pathways in plasma low-density lipoprotein-cholesterol lowering and gallstone disease. Eur J Epidemiol. 2024;39: 857–867. doi:10.1007/s10654-024-01141-5

26. Burgess S, Small DS, Thompson SG. A review of instrumental variable estimators for Mendelian randomization. Stat Methods Med Res. 2017;26: 2333–2355. doi:10.1177/0962280215597579

27. Yavorska OO, Burgess S. MendelianRandomization: an R package for performing Mendelian randomization analyses using summarized data. International Journal of Epidemiology. 2017;46: 1734–1739. doi:10.1093/ije/dyx034

28. Benjamini Y, Hochberg Y. Controlling the False Discovery Rate: A Practical and Powerful Approach to Multiple Testing. Journal of the Royal Statistical Society Series B: Statistical Methodology. 1995;57: 289–300. doi:10.1111/j.2517-6161.1995.tb02031.x

29. Hernán MA. The hazards of hazard ratios. Epidemiology. 2010;21: 13–15. doi:10.1097/EDE.0b013e3181c1ea43

30. Oliva M, Muñoz-Aguirre M, Kim-Hellmuth S, Wucher V, Gewirtz ADH, Cotter DJ, et al. The impact of sex on gene expression across human tissues. Science. 2020;369: eaba3066. doi:10.1126/science.aba3066

31. Jia F, Fei S-F, Tong D-B, Xue C, Li J-J. Sex difference in circulating PCSK9 and its clinical implications. Front Pharmacol. 2022;13: 953845. doi:10.3389/fphar.2022.953845

