## Supplemental Figures for "PCSK9 and breast cancer survival: a Mendelian Randomization study"

### MR of PCSK9 on Breast Cancer Survival, Pott et al. (2025)

**Figure S1: Study design flowchart**. We selected independent, genome-wide significant instruments for the (MV)MR-IVW approach (blue), and rs562556 with at least nominal significance for the MR-ratio approach (red) from three data sources: Pott et al. [15], GTEx v10 [17], and the GLGC [18,19]. The primary outcome of interest was breast cancer survival (BCS), using data from the Breast Cancer Association Consortium (BCAC) [20], FinnGen [21], or Mei et al. [14], and secondary outcomes were BC [21], coronary artery disease (CAD) [22] and parental age at death [23] as proxy for survival. The MR-ratio method could be applied to all outcomes, while the MR-IVW approach cannot be performed in Mei et al. as only data on rs562556 was provided.

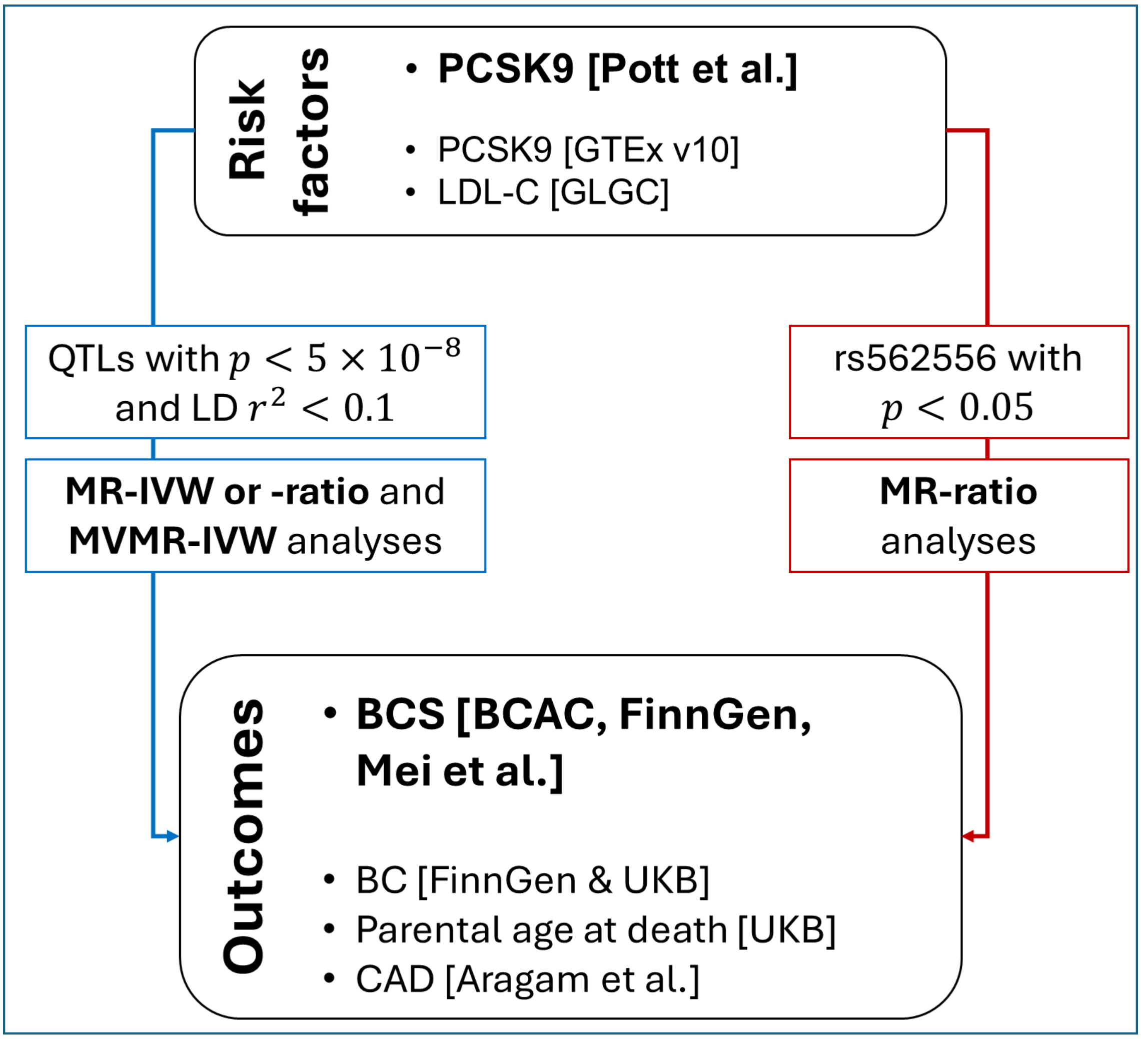

**Figure S2: Forest plot of the MR-ratio analyses of rs562556 on BC survival.** The log hazard ratios (logHR) for BC survival per 1 SD increment in PCSK9 levels are given for A) protein levels and B) gene expression (GE). The ratio estimates were not significantly different from 0 in when testing PCSK9 on the BCAC or FinnGen data, but significant when using data from the small studies reported by Mei et al. *[14]*. * significant after multiple testing correction.

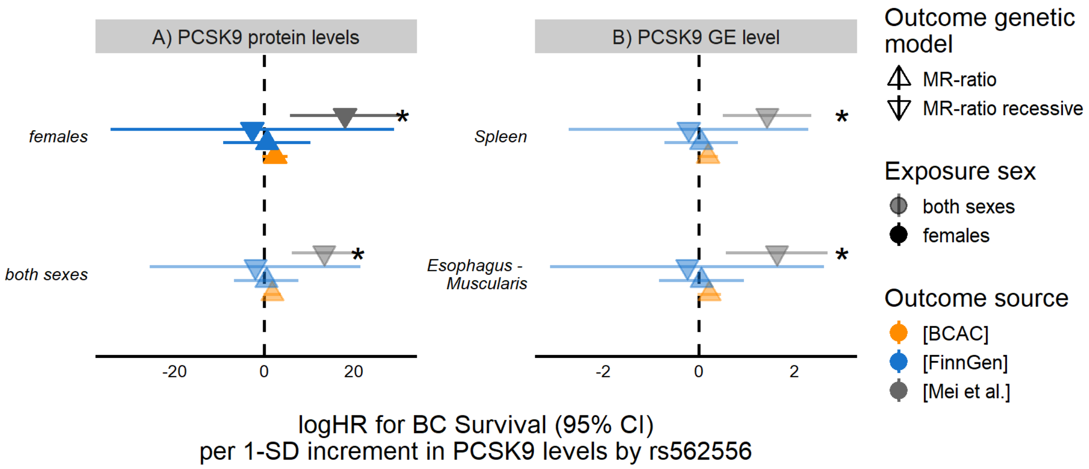

**Figure S3: Forest plot of the MR analyses on BC survival.** The log hazard ratios (logHR) for BC survival per 1 SD increment in PCSK9 levels are given for protein expression (PE) and selected tissues (gene expression, GE). For PE, the same four instruments were used for females and sex-combined exposure data. For GE, the best eQTLs per tissue were used, either with the ratio method (only one independent eQTL) or MR-IVW (multiple independent eQTLs).

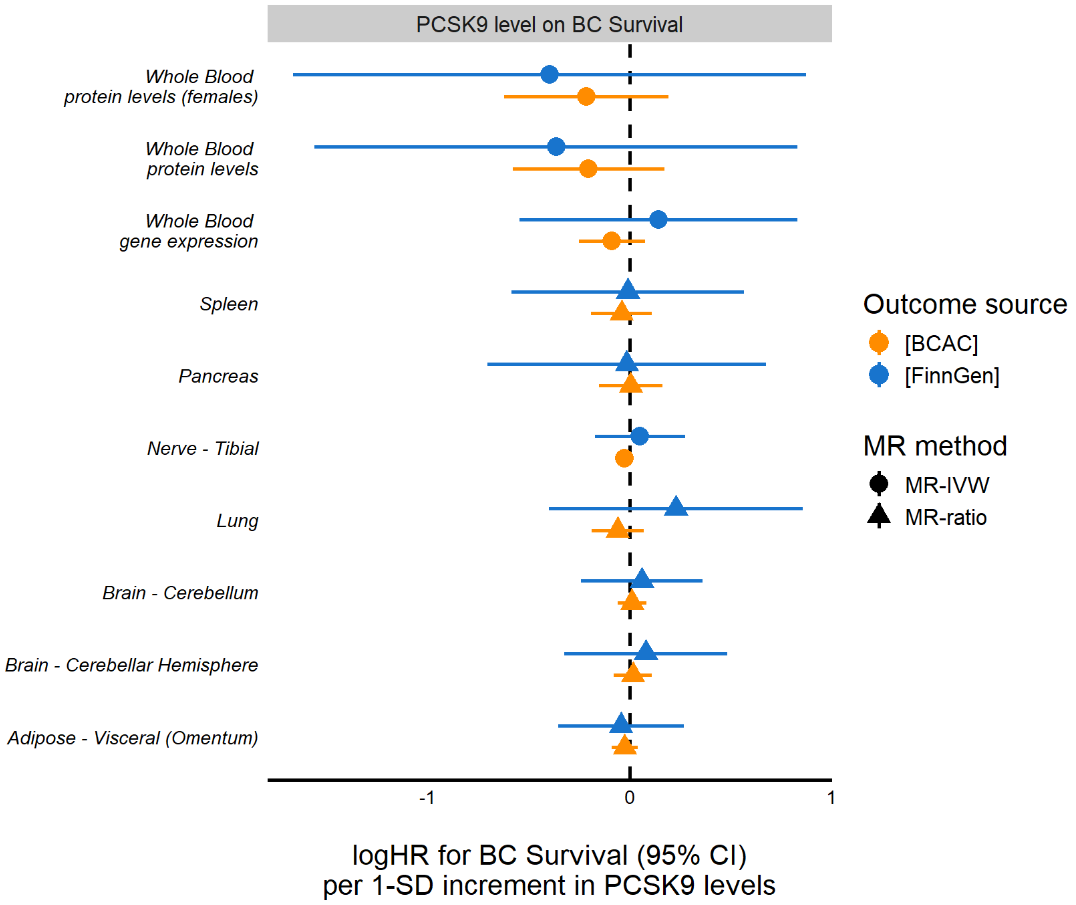

**Figure S4: Scatter plot of MR-IVW estimates using PCSK9 levels in females as exposure** and A) BC survival (from BCAC) B) BC incidence (from FinnGen + UKB meta-analysis) C) Parental age at death (from Pilling et al., UKB) and D) Coronary Artery Disease (CAD, from Aragam et al., females only) as outcome data. Error bars indicate the 95% confidence intervals of the SNP effects. The blue lines indicate the MR-IVW estimates, which were not significant for panel A and B (BC survival and incident, respectively), but significant for the positive controls in panel C and D (parental age at death and CAD in females, respectively). Estimates and F-statistics can be found in Supplemental Table S5.

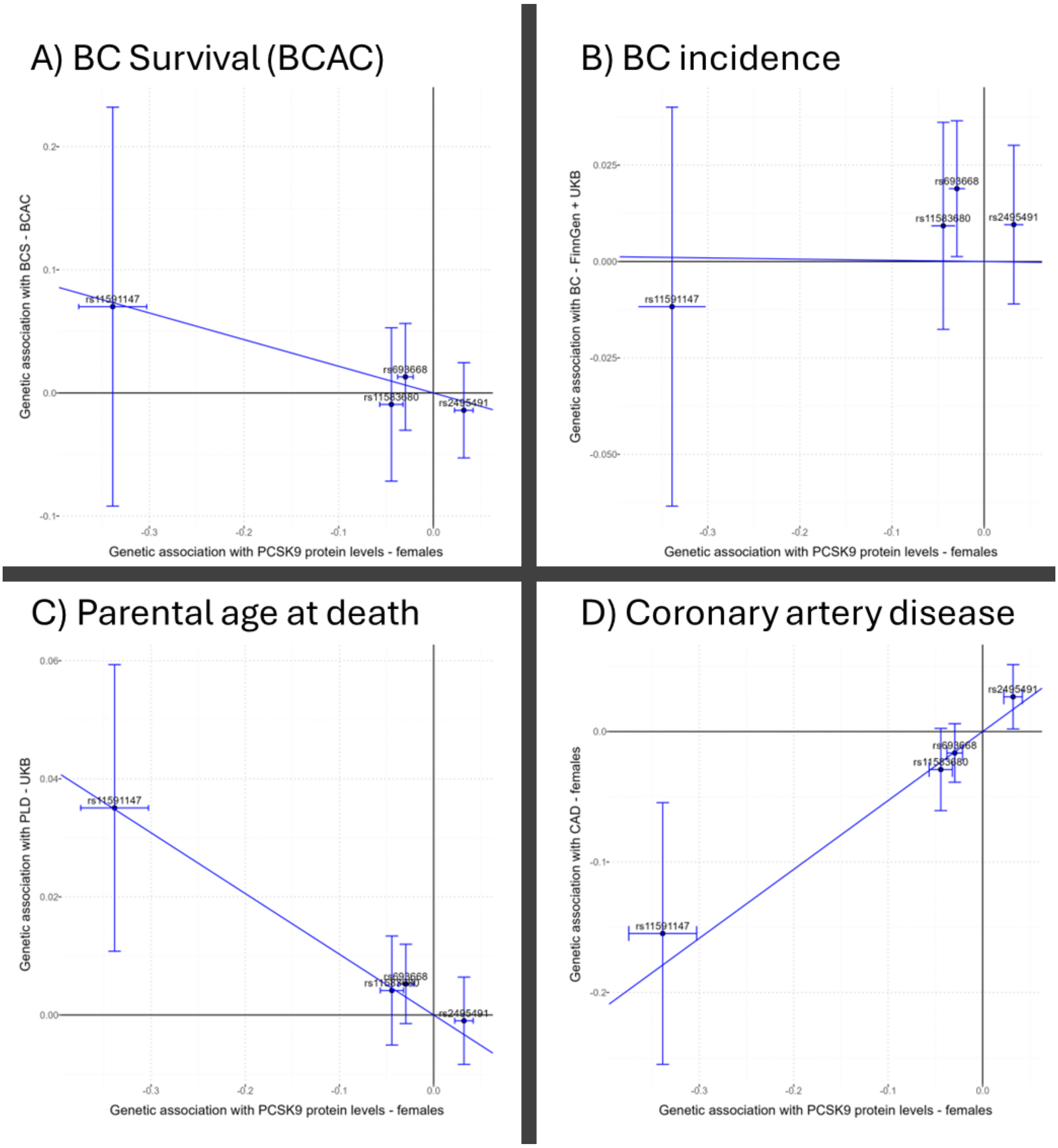

**Figure S5: Scatter plot of MR-IVW estimates using *HMGCR* instruments, LDL-C levels in exposure** and A) BC survival (from BCAC, sex-combined LDL-C data) B) BC survival (from BCAC, females only LDL-C data) C) Coronary Artery Disease (CAD, from Aragam et al., sex-combined exposure and outcome data) and D) CAD (from Aragam et al., females only exposure and outcome data) as outcome data. Error bars indicate the 95% confidence intervals of the SNP effects. The blue lines indicate the MR-IVW estimates, which were significant for all but panel D (CAD females). Estimates and F-statistics can be found in Supplemental Table S5.

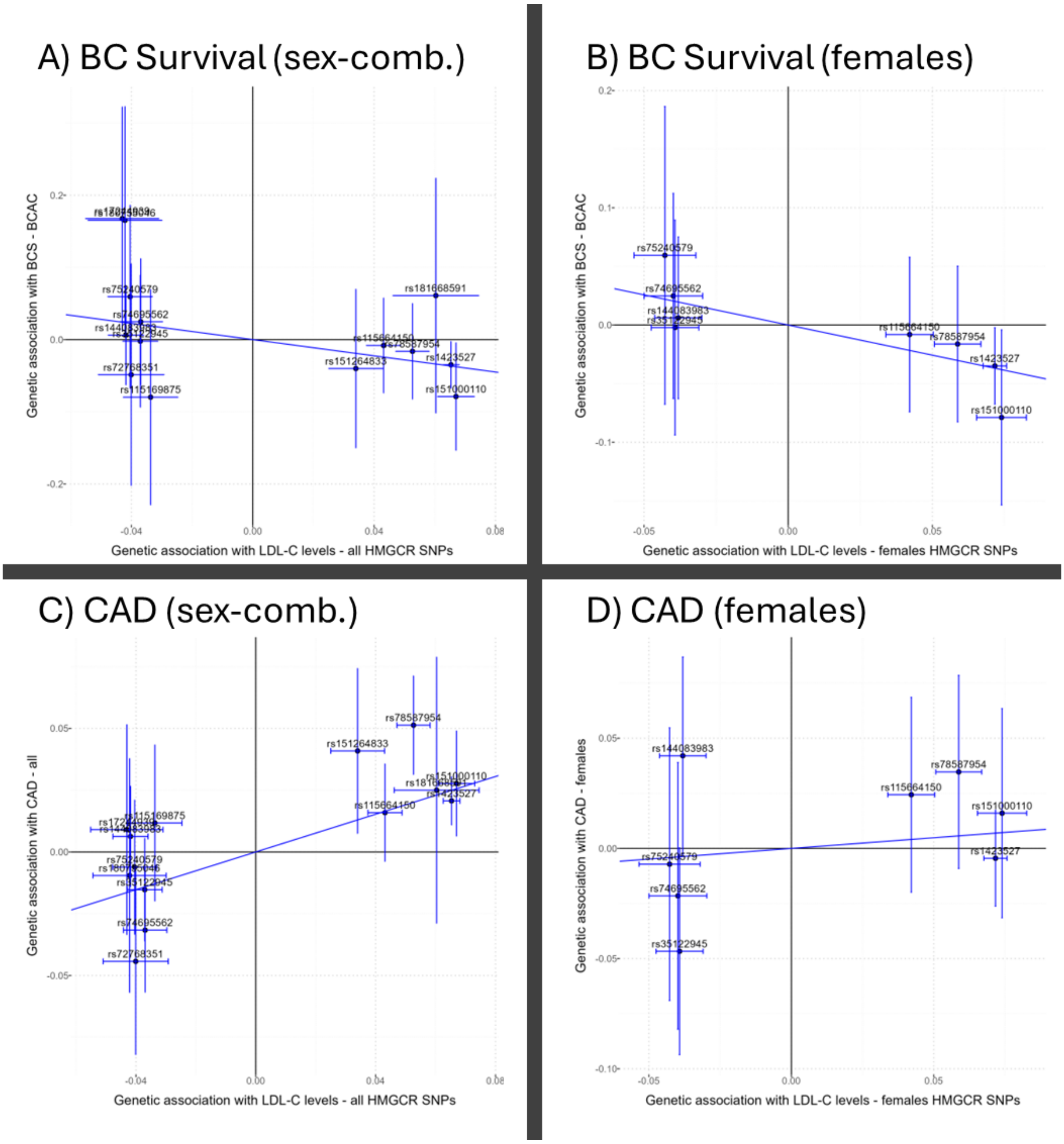

**Figure S6: Forest plot of the MR analyses on all other outcomes.** The log odds ratio (logOR) for BC or CAD risk, or the 1-SD increment for PAD per 1 SD increment in PCSK9 levels are given for protein expression (PE) and selected tissues (gene expression, GE). In the left panel, the MR-ratio results are shown, representing the functionality of PCSK9 to bind to LRP1. On the right panel, the MR-IVW results are shown, using the best QTLs per subgroup or tissue and hence representing the effect of PCSK9 levels. CAD served as positive control, and indeed the effect of PCSK9 on CAD was positive and significant using CAD sex-combined data. BC served as negative control, and indeed there was no significant effect detected in any test.

* significant after multiple testing correction; WB – whole blood; PE – protein expression; F – females only; CH – cerebellar hemisphere; BC – breast cancer; CAD – Coronary Artery Disease; PAD – Parental Age at Death

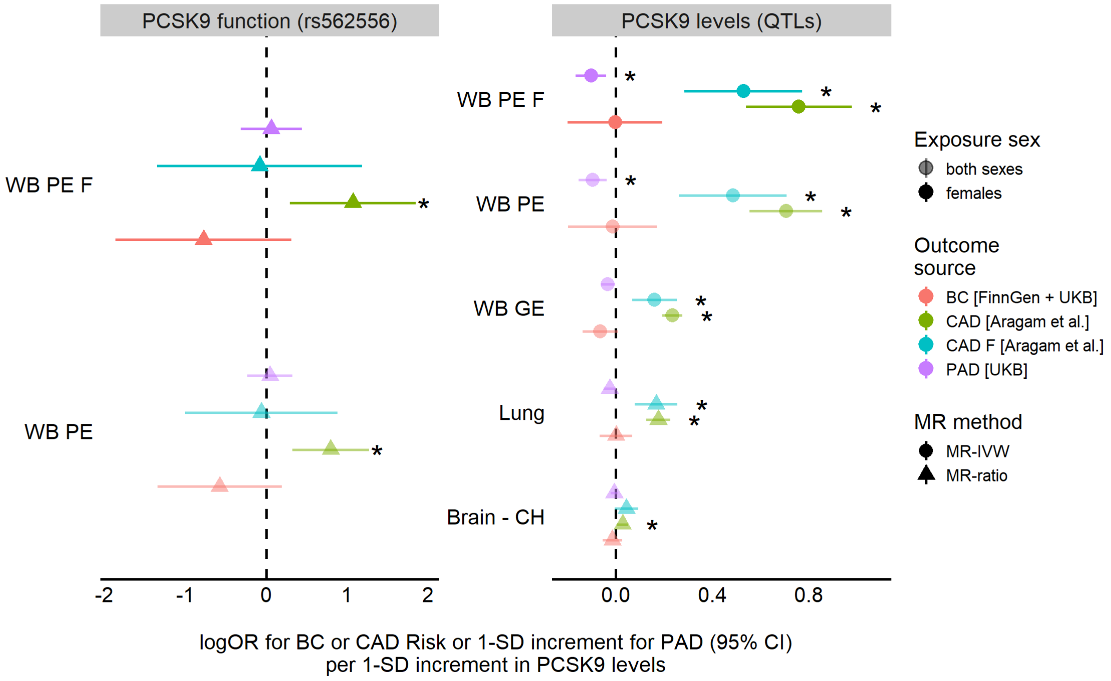

**Figure S7: Forest plots of the MR analyses of LDL-C and of the MVMR analyses of PCSK9 conditional on LDL-C on all other outcomes.** The log odds ratio (logOR) for BC or CAD risk, or the 1-SD increment for PAD per 1 SD increment in LDL-C or PCSK9 levels are given per selected instruments. **Left Panel**: As expected, LDL-C had a significant positive effect on CAD and a significant negative effect on PAD when using all instruments or instruments at *PCSK9*, while BC risk was not affected by it. When using instruments at *HMGCR* only, the effect was only significant in the sex-combined CAD outcome, while the BC risk effect was nominally significant, but not after correction for multiple testing. **Right panel**: PCSK9 was nominal significantly associated with CAD in the sex-combined setting, but not after multiple testing correction.

* significant after multiple testing correction; BC – breast cancer; CAD – Coronary Artery Disease; PAD – Parental Age at Death

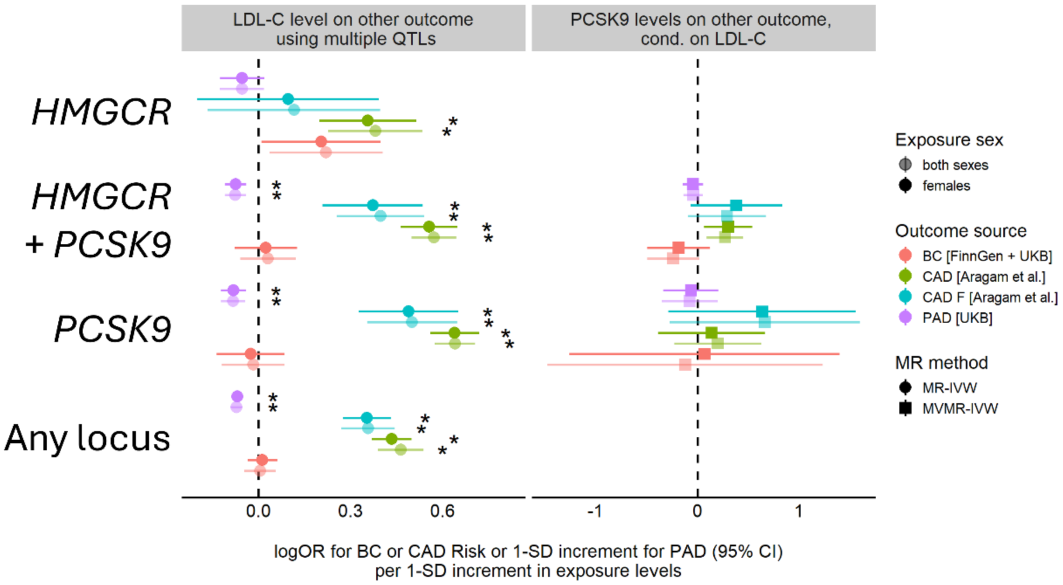
