## Supplemental Note for "PCSK9 and breast cancer survival: a Mendelian Randomization study"

MR of PCSK9 on Breast Cancer Survival, Pott et al. (2025)

### FinnGen cohort

Genotyping, imputation and QC

Illumina and Affymetrix DNA microarrays were used for genotyping. Genotype data were quality controlled to exclude variants with a low Hardy-Weinberg equilibrium (HWE) p-value ($<1x10^{-6}$), minor allele count (MAC) below three, and high missingness (cut-off 2%), as well as individuals with high genotype missingness (cut-off 5%), high levels of heterozygosity (±4 SD), non-Finnish ancestry, and individuals whose sex did not match the genotype data. In prephasing, Eagle 2.3.5 was utilised, with 20,000 serving as the conditioning haplotype threshold. Beagle 4.1 (version 08Jun17.d8b) was utilised for genotype imputation, and Finnish SISu v4.2 was utilised as a reference panel. The whole imputation protocol has been described at https://docs.finngen.fi/finngen-data-specifics/red-library-data-individual-level-data/genotype-data/imputation-panel/sisu-v4.2-reference-panel. For post-imputation quality control, we excluded variants with imputation information less than 0.6.

FinnGen R12 Ethics statement

Study subjects in FinnGen provided informed consent for biobank research, based on the Finnish Biobank Act. Alternatively, separate research cohorts, collected prior the Finnish Biobank Act came into effect (in September 2013) and start of FinnGen (August 2017), were collected based on study-specific consents and later transferred to the Finnish biobanks after approval by Fimea (Finnish Medicines Agency), the National Supervisory Authority for Welfare and Health. Recruitment protocols followed the biobank protocols approved by Fimea. The Coordinating Ethics Committee of the Hospital District of Helsinki and Uusimaa (HUS) statement number for the FinnGen study is Nr HUS/990/2017.

The FinnGen study is approved by Finnish Institute for Health and Welfare (permit numbers: THL/2031/6.02.00/2017, THL/1101/5.05.00/2017, THL/341/6.02.00/2018, THL/2222/6.02.00/2018, THL/283/6.02.00/2019, THL/1721/5.05.00/2019 and THL/1524/5.05.00/2020), Digital and population data service agency (permit numbers: VRK43431/2017-3, VRK/6909/2018-3, VRK/4415/2019-3), the Social Insurance Institution (permit numbers: KELA 58/522/2017, KELA 131/522/2018, KELA 70/522/2019, KELA 98/522/2019, KELA 134/522/2019, KELA 138/522/2019, KELA 2/522/2020, KELA 16/522/2020), Findata permit numbers THL/2364/14.02/2020, THL/4055/14.06.00/2020, THL/3433/14.06.00/2020, THL/4432/14.06/2020, THL/5189/14.06/2020, THL/5894/14.06.00/2020, THL/6619/14.06.00/2020, THL/209/14.06.00/2021, THL/688/14.06.00/2021, THL/1284/14.06.00/2021, THL/1965/14.06.00/2021, THL/5546/14.02.00/2020, THL/2658/14.06.00/2021, THL/4235/14.06.00/2021, Statistics Finland (permit numbers: TK-53-1041-17 and TK/143/07.03.00/2020 (earlier TK-53-90-20) TK/1735/07.03.00/2021, TK/3112/07.03.00/2021) and Finnish Registry for Kidney Diseases permission/extract from the meeting minutes on 4th July 2019.

The Biobank Access Decisions for FinnGen samples and data utilized in FinnGen Data Freeze 12 include: THL Biobank BB2017_55, BB2017_111, BB2018_19, BB_2018_34, BB_2018_67, BB2018_71, BB2019_7, BB2019_8, BB2019_26, BB2020_1, BB2021_65, Finnish Red Cross Blood Service Biobank 7.12.2017, Helsinki Biobank HUS/359/2017, HUS/248/2020, HUS/430/2021 §28, §29, HUS/150/2022 §12, §13, §14, §15, §16, §17, §18, §23, §58, §59, HUS/128/2023 §18, Auria Biobank AB17-5154 and amendment #1 (August 17 2020) and amendments BB_2021-0140, BB_2021-0156 (August 26 2021, Feb 2 2022), BB_2021-0169, BB_2021-0179, BB_2021-0161, AB20-5926 and amendment #1 (April 23 2020) and it´s modifications (Sep 22 2021), BB_2022-0262, BB_2022-0256, Biobank Borealis of Northern Finland_2017_1013, 2021_5010, 2021_5010 Amendment, 2021_5018, 2021_5018 Amendment, 2021_5015, 2021_5015 Amendment, 2021_5015 Amendment_2, 2021_5023, 2021_5023 Amendment, 2021_5023 Amendment_2, 2021_5017, 2021_5017 Amendment, 2022_6001, 2022_6001 Amendment, 2022_6006 Amendment, 2022_6006 Amendment, 2022_6006 Amendment_2, BB22-0067, 2022_0262, 2022_0262 Amendment, Biobank of Eastern Finland 1186/2018 and amendment 22§/2020, 53§/2021, 13§/2022, 14§/2022, 15§/2022, 27§/2022, 28§/2022, 29§/2022, 33§/2022, 35§/2022, 36§/2022, 37§/2022, 39§/2022, 7§/2023, 32§/2023, 33§/2023, 34§/2023, 35§/2023, 36§/2023, 37§/2023, 38§/2023, 39§/2023, 40§/2023, 41§/2023, Finnish Clinical Biobank Tampere MH0004 and amendments (21.02.2020 & 06.10.2020), BB2021-0140 8§/2021, 9§/2021, §9/2022, §10/2022, §12/2022, 13§/2022, §20/2022, §21/2022, §22/2022, §23/2022, 28§/2022, 29§/2022, 30§/2022, 31§/2022, 32§/2022, 38§/2022, 40§/2022, 42§/2022, 1§/2023, Central Finland Biobank 1-2017, BB_2021-0161, BB_2021-0169, BB_2021-0179, BB_2021-0170, BB_2022-0256, BB_2022-0262, BB22-0067, Decision allowing to continue data processing until 31st Aug 2024 for projects: BB_2021-0179, BB22-0067,BB_2022-0262, BB_2021-0170, BB_2021-0164, BB_2021-0161, and BB_2021-0169, and Terveystalo Biobank STB 2018001 and amendment 25th Aug 2020, Finnish Hematological Registry and Clinical Biobank decision 18th June 2021, Arctic biobank P0844: ARC_2021_1001.

FinnGen authors and their affiliations

| Full Name | Affiliation | Role 1 | Role 2 |
|---|---|---|---|
| **Aarno Palotie** | Institute for Molecular Medicine Finland (FIMM), HiLIFE, University of Helsinki, Helsinki, Finland; Broad Institute of MIT and Harvard; Massachusetts General Hospital | Steering Committee | Steering Committee |
| **Mark Daly** | Institute for Molecular Medicine Finland (FIMM), HiLIFE, University of Helsinki, Helsinki, Finland; Broad Institute of MIT and Harvard; Massachusetts General Hospital | Steering Committee | Steering Committee |
| **Bridget Riley-Gills** | Abbvie, Chicago, IL, United States | Steering Committee | Pharmaceutical companies |
| **Howard Jacob** | Abbvie, Chicago, IL, United States | Steering Committee | Pharmaceutical companies |
| **Coralie Viollet** | Astra Zeneca, Cambridge, United Kingdom | Steering Committee | Pharmaceutical companies |
| **Slavé Petrovski** | Astra Zeneca, Cambridge, United Kingdom | Steering Committee | Pharmaceutical companies |
| **Alix Berton** | Bayer AG, Leverkusen, Germany | Steering Committee | Pharmaceutical companies |
| **Santha Ramakrishnan** | Bayer AG, Leverkusen, Germany | Steering Committee | Pharmaceutical companies |
| **Ellen Tsai** | Biogen, Cambridge, MA, United States | Steering Committee | Pharmaceutical companies |
| **George Okafo** | Boehringer Ingelheim, Ingelheim am Rhein, Germany | Steering Committee | Pharmaceutical companies |
| **Emily Holzinger** | Bristol Myers Squibb, New York, NY, United States | Steering Committee | Pharmaceutical companies |
| **Robert Plenge** | Bristol Myers Squibb, New York, NY, United States | Steering Committee | Pharmaceutical companies |

| | | | |
|---|---|---|---|
| **Joseph Maranville** | Bristol Myers Squibb, New York, NY, United States | Steering Committee | Pharmaceutical companies |
| **Mark McCarthy** | Genentech, San Francisco, CA, United States | Steering Committee | Pharmaceutical companies |
| **Rion Pendergrass** | Genentech, San Francisco, CA, United States | Steering Committee | Pharmaceutical companies |
| **Jonathan Davitte** | GlaxoSmithKline, Collegeville, PA, United States | Steering Committee | Pharmaceutical companies |
| **Kirsi Auro** | GlaxoSmithKline, Espoo, Finland | Steering Committee | Pharmaceutical companies |
| **Simonne Longerich** | Merck, Kenilworth, NJ, United States | Steering Committee | Pharmaceutical companies |
| **Anders Mälarstig** | Pfizer, New York, NY, United States | Steering Committee | Pharmaceutical companies |
| **Anna Vlahiotis** | Pfizer, New York, NY, United States | Steering Committee | Pharmaceutical companies |
| **Katherine Klinger** | Translational Sciences, Sanofi R&D, Framingham, MA, USA | Steering Committee | Pharmaceutical companies |
| **Clement Chatelain** | Translational Sciences, Sanofi R&D, Framingham, MA, USA | Steering Committee | Pharmaceutical companies |
| **Jorg Blankenstein** | Translational Sciences, Sanofi R&D, Framingham, MA, USA | Steering Committee | Pharmaceutical companies |
| **Karol Estrada** | Maze Therapeutics, San Francisco, CA, United States | Steering Committee | Pharmaceutical companies |
| **Robert Graham** | Maze Therapeutics, San Francisco, CA, United States | Steering Committee | Pharmaceutical companies |
| **Dawn Waterworth** | Johnson & Johnson Innovative Medicine, Spring House, PA, United States | Steering Committee | Pharmaceutical companies |
| **Chris O´Donnell** | Novartis Institutes for BioMedical Research, Cambridge, MA, United States | Steering Committee | Pharmaceutical companies |
| **Nicole Renaud** | Novartis Institutes for BioMedical Research, Cambridge, MA, United States | Steering Committee | Pharmaceutical companies |
| **Tomi P. Mäkelä** | HiLIFE, University of Helsinki, Finland, Finland | Steering Committee | University of Helsinki & Biobanks |
| **Jaakko Kaprio** | Institute for Molecular Medicine Finland (FIMM), HiLIFE, University of Helsinki, Helsinki, Finland | Steering Committee | University of Helsinki & Biobanks |
| **Minna Ruddock** | Arctic biobank / University of Oulu | Steering Committee | University of Helsinki & Biobanks |
| **Petri Virolainen** | Auria Biobank / University of Turku / Wellbeing Services County of Southwest Finland, Turku, Finland | Steering Committee | University of Helsinki & Biobanks |
| **Antti Hakanen** | Auria Biobank / University of Turku / Wellbeing Services County of Southwest Finland, Turku, Finland | Steering Committee | University of Helsinki & Biobanks |
| **Terhi Kilpi** | THL Biobank / Finnish Institute for Health and Welfare (THL), Helsinki, Finland | Steering Committee | University of Helsinki & Biobanks |
| **Markus Perola** | THL Biobank / Finnish Institute for Health and Welfare (THL), Helsinki, Finland | Steering Committee | University of Helsinki & Biobanks |
| **Jukka Partanen** | Finnish Red Cross Blood Service / Finnish Hematology Registry and Clinical Biobank, Helsinki, Finland | Steering Committee | University of Helsinki & Biobanks |
| **Taneli Raivio** | Helsinki Biobank / Helsinki University and Hospital District of Helsinki and Uusimaa, Helsinki | Steering Committee | University of Helsinki & Biobanks |
| **Raisa Serpi** | Northern Finland Biobank Borealis / University of Oulu / Wellbeing services county of North Ostrobothnia, Oulu, Finland | Steering Committee | University of Helsinki & Biobanks |
| **Kati Kristiansson** | Finnish Clinical Biobank Tampere / University of Tampere / Wellbeing Services County of Pirkanmaa, Tampere, Finland | Steering Committee | University of Helsinki & Biobanks |
| **Veli-Matti Kosma** | Biobank of Eastern Finland / University of Eastern Finland / Wellbeing services county of North Savo, Kuopio, Finland | Steering Committee | University of Helsinki & Biobanks |

| | | | |
|---|---|---|---|
| **Jari Laukkanen** | Central Finland Biobank / University of Jyväskylä / Wellbeing Services County of Central Finland, Jyväskylä, Finland | Steering Committee | University of Helsinki & Biobanks |
| **Tom Southerington** | Finnish Biobank Cooperative – FINBB | Steering Committee | University of Helsinki & Biobanks |
| **Outi Tuovila** | Business Finland, Helsinki, Finland | Steering Committee | Other Experts/ Non-Voting Members |
| **Jeffrey Waring** | Abbvie, Chicago, IL, United States | Scientific Committee | Pharmaceutical companies |
| **Bridget Riley-Gillis** | Abbvie, Chicago, IL, United States | Scientific Committee | Pharmaceutical companies |
| **Fedik Rahimov** | Abbvie, Chicago, IL, United States | Scientific Committee | Pharmaceutical companies |
| **Ioanna Tachmazidou** | Astra Zeneca, Cambridge, United Kingdom | Scientific Committee | Pharmaceutical companies |
| **Alix Berton** | Bayer AG, Leverkusen, Germany | Scientific Committee | Pharmaceutical companies |
| **Santha Ramakrishnan** | Bayer AG, Leverkusen, Germany | Scientific Committee | Pharmaceutical companies |
| **Ellen Tsai** | Biogen, Cambridge, MA, United States | Scientific Committee | Pharmaceutical companies |
| **Zhihao Ding** | Boehringer Ingelheim, Ingelheim am Rhein, Germany | Scientific Committee | Pharmaceutical companies |
| **Marc Jung** | Boehringer Ingelheim, Ingelheim am Rhein, Germany | Scientific Committee | Pharmaceutical companies |
| **Hanati Tuoken** | Boehringer Ingelheim, Ingelheim am Rhein, Germany | Scientific Committee | Pharmaceutical companies |
| **Shameek Biswas** | Bristol Myers Squibb, New York, NY, United States | Scientific Committee | Pharmaceutical companies |
| **Benjamin Sun** | Bristol Myers Squibb, New York, NY, United States | Scientific Committee | Pharmaceutical companies |
| **Rion Pendergrass** | Genentech, San Francisco, CA, United States | Scientific Committee | Pharmaceutical companies |
| **Jonathan Davitte** | GlaxoSmithKline, Collegeville, PA, United States | Scientific Committee | Pharmaceutical companies |
| **Neha Raghavan** | Merck, Kenilworth, NJ, United States | Scientific Committee | Pharmaceutical companies |
| **Adriana Huertas-Vazquez** | Merck, Kenilworth, NJ, United States | Scientific Committee | Pharmaceutical companies |
| **Jae-Hoon Sul** | Merck, Kenilworth, NJ, United States | Scientific Committee | Pharmaceutical companies |
| **Anders Mälarstig** | Pfizer, New York, NY, United States | Scientific Committee | Pharmaceutical companies |
| **Xinli Hu** | Pfizer, New York, NY, United States | Scientific Committee | Pharmaceutical companies |
| **Åsa Hedman** | Pfizer, New York, NY, United States | Scientific Committee | Pharmaceutical companies |
| **Katherine Klinger** | Translational Sciences, Sanofi R&D, Framingham, MA, USA | Scientific Committee | Pharmaceutical companies |
| **Robert Graham** | Maze Therapeutics, San Francisco, CA, United States | Scientific Committee | Pharmaceutical companies |
| **Dawn Waterworth** | Johnson & Johnson Innovative Medicine, Spring House, PA, United States | Scientific Committee | Pharmaceutical companies |
| **Nicole Renaud** | Novartis Institutes for BioMedical Research, Cambridge, MA, United States | Scientific Committee | Pharmaceutical companies |
| **Ma´en Obeidat** | Novartis Institutes for BioMedical Research, Cambridge, MA, United States | Scientific Committee | Pharmaceutical companies |
| **Jonathan Chung** | Novartis Institutes for BioMedical Research, Cambridge, MA, United States | Scientific Committee | Pharmaceutical companies |
| **Jonas Zierer** | Novartis Institutes for BioMedical Research, Cambridge, MA, United States | Scientific Committee | Pharmaceutical companies |
| **Mari Niemi** | Novartis Institutes for BioMedical Research, Cambridge, MA, United States | Scientific Committee | Pharmaceutical companies |
| **Samuli Ripatti** | Institute for Molecular Medicine Finland (FIMM), HiLIFE, University of Helsinki, Helsinki, Finland | Scientific Committee | University of Helsinki & Biobanks |
| **Johanna Schleutker** | Auria Biobank / University of Turku / Wellbeing Services County of Southwest Finland, Turku, Finland | Scientific Committee | University of Helsinki & Biobanks |
| **Markus Perola** | THL Biobank / Finnish Institute for Health and Welfare (THL), Helsinki, Finland | Scientific Committee | University of Helsinki & Biobanks |

| | | | |
|---|---|---|---|
| **Mikko Arvas** | Finnish Red Cross Blood Service / Finnish Hematology Registry and Clinical Biobank, Helsinki, Finland | Scientific Committee | University of Helsinki & Biobanks |
| **Olli Carpén** | Helsinki Biobank / Helsinki University and Hospital District of Helsinki and Uusimaa, Helsinki | Scientific Committee | University of Helsinki & Biobanks |
| **Reetta Hinttala** | Northern Finland Biobank Borealis / University of Oulu / Wellbeing services county of North Ostrobothnia, Oulu, Finland | Scientific Committee | University of Helsinki & Biobanks |
| **Johannes Kettunen** | Northern Finland Biobank Borealis / University of Oulu / Wellbeing services county of North Ostrobothnia, Oulu, Finland | Scientific Committee | University of Helsinki & Biobanks |
| **Arto Mannermaa** | Biobank of Eastern Finland / University of Eastern Finland / Wellbeing services county of North Savo, Kuopio, Finland | Scientific Committee | University of Helsinki & Biobanks |
| **Katriina Aalto-Setälä** | Faculty of Medicine and Health Technology, Tampere University, Tampere, Finland | Scientific Committee | University of Helsinki & Biobanks |
| **Mika Kähönen** | Finnish Clinical Biobank Tampere / University of Tampere / Wellbeing Services County of Pirkanmaa, Tampere, Finland | Scientific Committee | University of Helsinki & Biobanks |
| **Jari Laukkanen** | Central Finland Biobank / University of Jyväskylä / Wellbeing Services County of Central Finland, Jyväskylä, Finland | Scientific Committee | University of Helsinki & Biobanks |
| **Johanna Mäkelä** | FINBB - Finnish biobank cooperative | Scientific Committee | University of Helsinki & Biobanks |
| **Reetta Kälviäinen** | Wellbeing services county of North Savo, Kuopio, Finland | Clinical Groups | Neurology Group |
| **Valtteri Julkunen** | Wellbeing services county of North Savo, Kuopio, Finland | Clinical Groups | Neurology Group |
| **Hilkka Soininen** | Wellbeing services county of North Savo, Kuopio, Finland | Clinical Groups | Neurology Group |
| **Anne Remes** | Wellbeing services county of North Ostrobothnia, Oulu, Finland | Clinical Groups | Neurology Group |
| **Mikko Hiltunen** | University of Eastern Finland, Kuopio, Finland | Clinical Groups | Neurology Group |
| **Jukka Peltola** | Wellbeing Services County of Pirkanmaa, Tampere, Finland | Clinical Groups | Neurology Group |
| **Minna Raivio** | Hospital District of Helsinki and Uusimaa, Helsinki, Finland | Clinical Groups | Neurology Group |
| **Pentti Tienari** | Hospital District of Helsinki and Uusimaa, Helsinki, Finland | Clinical Groups | Neurology Group |
| **Juha Rinne** | Wellbeing Services County of Southwest Finland, Turku, Finland | Clinical Groups | Neurology Group |
| **Roosa Kallionpää** | Wellbeing Services County of Southwest Finland, Turku, Finland | Clinical Groups | Neurology Group |
| **Juulia Partanen** | Institute for Molecular Medicine Finland, HiLIFE, University of Helsinki, Finland | Clinical Groups | Neurology Group |
| **Adam Ziemann** | Abbvie, Chicago, IL, United States | Clinical Groups | Neurology Group |
| **Nizar Smaoui** | Abbvie, Chicago, IL, United States | Clinical Groups | Neurology Group |
| **Anne Lehtonen** | Abbvie, Chicago, IL, United States | Clinical Groups | Neurology Group |
| **Susan Eaton** | Biogen, Cambridge, MA, United States | Clinical Groups | Neurology Group |
| **Shameek Biswas** | Bristol Myers Squibb, New York, NY, United States | Clinical Groups | Neurology Group |
| **Natalie Bowers** | Genentech, San Francisco, CA, United States | Clinical Groups | Neurology Group |
| **Edmond Teng** | Genentech, San Francisco, CA, United States | Clinical Groups | Neurology Group |
| **Rion Pendergrass** | Genentech, San Francisco, CA, United States | Clinical Groups | Neurology Group |
| **Fanli Xu** | GlaxoSmithKline, Brentford, United Kingdom | Clinical Groups | Neurology Group |

| | | | |
|---|---|---|---|
| **David Pulford** | GlaxoSmithKline, Stevenage, United Kingdom | Clinical Groups | Neurology Group |
| **Kirsi Auro** | GlaxoSmithKline, Espoo, Finland | Clinical Groups | Neurology Group |
| **Laura Addis** | GlaxoSmithKline, Brentford, United Kingdom | Clinical Groups | Neurology Group |
| **John Eicher** | GlaxoSmithKline, Brentford, United Kingdom | Clinical Groups | Neurology Group |
| **Qingqin S Li** | Johnson & Johnson Innovative Medicine, Titusville, NJ 08560, United States | Clinical Groups | Neurology Group |
| **Karen He** | Johnson & Johnson Innovative Medicine, Spring House, PA, United States | Clinical Groups | Neurology Group |
| **Ekaterina Khramtsova** | Johnson & Johnson Innovative Medicine, Spring House, PA, United States | Clinical Groups | Neurology Group |
| **Neha Raghavan** | Merck, Kenilworth, NJ, United States | Clinical Groups | Neurology Group |
| **Martti Färkkilä** | Hospital District of Helsinki and Uusimaa, Helsinki, Finland | Clinical Groups | Gastroenterology Group |
| **Jukka Koskela** | Hospital District of Helsinki and Uusimaa, Helsinki, Finland | Clinical Groups | Gastroenterology Group |
| **Sampsa Pikkarainen** | Hospital District of Helsinki and Uusimaa, Helsinki, Finland | Clinical Groups | Gastroenterology Group |
| **Airi Jussila** | Wellbeing Services County of Pirkanmaa, Tampere, Finland | Clinical Groups | Gastroenterology Group |
| **Katri Kaukinen** | Wellbeing Services County of Pirkanmaa, Tampere, Finland | Clinical Groups | Gastroenterology Group |
| **Timo Blomster** | Wellbeing services county of North Ostrobothnia, Oulu, Finland | Clinical Groups | Gastroenterology Group |
| **Mikko Kiviniemi** | Wellbeing services county of North Savo, Kuopio, Finland | Clinical Groups | Gastroenterology Group |
| **Markku Voutilainen** | Wellbeing Services County of Southwest Finland, Turku, Finland | Clinical Groups | Gastroenterology Group |
| **Mark Daly** | Institute for Molecular Medicine, Finland (FIMM), HiLIFE, University of Helsinki, Helsinki, Finland; Broad Institute of MIT and Harvard; Massachusetts General Hospital | Clinical Groups | Gastroenterology Group |
| **Jeffrey Waring** | Abbvie, Chicago, IL, United States | Clinical Groups | Gastroenterology Group |
| **Nizar Smaoui** | Abbvie, Chicago, IL, United States | Clinical Groups | Gastroenterology Group |
| **Fedik Rahimov** | Abbvie, Chicago, IL, United States | Clinical Groups | Gastroenterology Group |
| **Anne Lehtonen** | Abbvie, Chicago, IL, United States | Clinical Groups | Gastroenterology Group |
| **Tim Lu** | Genentech, San Francisco, CA, United States | Clinical Groups | Gastroenterology Group |
| **Natalie Bowers** | Genentech, San Francisco, CA, United States | Clinical Groups | Gastroenterology Group |
| **Rion Pendergrass** | Genentech, San Francisco, CA, United States | Clinical Groups | Gastroenterology Group |
| **Linda McCarthy** | GlaxoSmithKline, Brentford, United Kingdom | Clinical Groups | Gastroenterology Group |
| **Amy Hart** | Johnson & Johnson Innovative Medicine, Spring House, PA, United States | Clinical Groups | Gastroenterology Group |
| **Meijian Guan** | Johnson & Johnson Innovative Medicine, Spring House, PA, United States | Clinical Groups | Gastroenterology Group |
| **Jason Miller** | Merck, Kenilworth, NJ, United States | Clinical Groups | Gastroenterology Group |
| **Kirsi Kalpala** | Pfizer, New York, NY, United States | Clinical Groups | Gastroenterology Group |
| **Melissa Miller** | Pfizer, New York, NY, United States | Clinical Groups | Gastroenterology Group |
| **Xinli Hu** | Pfizer, New York, NY, United States | Clinical Groups | Gastroenterology Group |
| **Kari Eklund** | Hospital District of Helsinki and Uusimaa, Helsinki, Finland | Clinical Groups | Rheumatology Group |
| **Antti Palomäki** | Wellbeing Services County of Southwest Finland, Turku, Finland | Clinical Groups | Rheumatology Group |
| **Pia Isomäki** | Wellbeing Services County of Pirkanmaa, Tampere, Finland | Clinical Groups | Rheumatology Group |
| **Laura Pirilä** | Wellbeing Services County of Southwest Finland, Turku, Finland | Clinical Groups | Rheumatology Group |

| | | | |
|---|---|---|---|
| **Oili Kaipiainen-Seppänen** | Wellbeing services county of North Savo, Kuopio, Finland | Clinical Groups | Rheumatology Group |
| **Johanna Huhtakangas** | Wellbeing services county of North Ostrobothnia, Oulu, Finland | Clinical Groups | Rheumatology Group |
| **Nina Mars** | Institute for Molecular Medicine Finland (FIMM), HiLIFE, University of Helsinki, Helsinki, Finland | Clinical Groups | Rheumatology Group |
| **Jeffrey Waring** | Abbvie, Chicago, IL, United States | Clinical Groups | Rheumatology Group |
| **Fedik Rahimov** | Abbvie, Chicago, IL, United States | Clinical Groups | Rheumatology Group |
| **Apinya Lertratanakul** | Abbvie, Chicago, IL, United States | Clinical Groups | Rheumatology Group |
| **Nizar Smaoui** | Abbvie, Chicago, IL, United States | Clinical Groups | Rheumatology Group |
| **Anne Lehtonen** | Abbvie, Chicago, IL, United States | Clinical Groups | Rheumatology Group |
| **Coralie Viollet** | AstraZeneca, Cambridge, United Kingdom | Clinical Groups | Rheumatology Group |
| **Marla Hochfeld** | Bristol Myers Squibb, New York, NY, United States | Clinical Groups | Rheumatology Group |
| **Natalie Bowers** | Genentech, San Francisco, CA, United States | Clinical Groups | Rheumatology Group |
| **Rion Pendergrass** | Genentech, San Francisco, CA, United States | Clinical Groups | Rheumatology Group |
| **Jorge Esparza Gordillo** | GlaxoSmithKline, Brentford, United Kingdom | Clinical Groups | Rheumatology Group |
| **Kirsi Auro** | GlaxoSmithKline, Espoo, Finland | Clinical Groups | Rheumatology Group |
| **Dawn Waterworth** | Johnson & Johnson Innovative Medicine, Spring House, PA, United States | Clinical Groups | Rheumatology Group |
| **Fabiana Farias** | Merck, Kenilworth, NJ, United States | Clinical Groups | Rheumatology Group |
| **Kirsi Kalpala** | Pfizer, New York, NY, United States | Clinical Groups | Rheumatology Group |
| **Nan Bing** | Pfizer, New York, NY, United States | Clinical Groups | Rheumatology Group |
| **Xinli Hu** | Pfizer, New York, NY, United States | Clinical Groups | Rheumatology Group |
| **Tarja Laitinen** | Wellbeing Services County of Pirkanmaa, Tampere, Finland | Clinical Groups | Pulmonology Group |
| **Margit Pelkonen** | Wellbeing services county of North Savo, Kuopio, Finland | Clinical Groups | Pulmonology Group |
| **Paula Kauppi** | Hospital District of Helsinki and Uusimaa, Helsinki, Finland | Clinical Groups | Pulmonology Group |
| **Hannu Kankaanranta** | University of Gothenburg, Gothenburg, Sweden/ Seinäjoki Central Hospital, Seinäjoki, Finland/ Tampere University, Tampere, Finland | Clinical Groups | Pulmonology Group |
| **Terttu Harju** | Wellbeing services county of North Ostrobothnia, Oulu, Finland | Clinical Groups | Pulmonology Group |
| **Riitta Lahesmaa** | Wellbeing Services County of Southwest Finland, Turku, Finland | Clinical Groups | Pulmonology Group |
| **Nizar Smaoui** | Abbvie, Chicago, IL, United States | Clinical Groups | Pulmonology Group |
| **Coralie Viollet** | AstraZeneca, Cambridge, United Kingdom | Clinical Groups | Pulmonology Group |
| **Susan Eaton** | Biogen, Cambridge, MA, United States | Clinical Groups | Pulmonology Group |
| **Hubert Chen** | Genentech, San Francisco, CA, United States | Clinical Groups | Pulmonology Group |
| **Rion Pendergrass** | Genentech, San Francisco, CA, United States | Clinical Groups | Pulmonology Group |
| **Natalie Bowers** | Genentech, San Francisco, CA, United States | Clinical Groups | Pulmonology Group |
| **Joanna Betts** | GlaxoSmithKline, Brentford, United Kingdom | Clinical Groups | Pulmonology Group |
| **Kirsi Auro** | GlaxoSmithKline, Espoo, Finland | Clinical Groups | Pulmonology Group |
| **Rajashree Mishra** | GlaxoSmithKline, Brentford, United Kingdom | Clinical Groups | Pulmonology Group |
| **Majd Mouded** | Novartis, Basel, Switzerland | Clinical Groups | Pulmonology Group |
| **Debby Ngo** | Novartis, Basel, Switzerland | Clinical Groups | Pulmonology Group |
| **Teemu Niiranen** | Finnish Institute for Health and Welfare (THL), Helsinki, Finland | Clinical Groups | Cardiometabolic Diseases Group |
| **Felix Vaura** | Finnish Institute for Health and Welfare (THL), Helsinki, Finland | Clinical Groups | Cardiometabolic Diseases Group |
| **Veikko Salomaa** | Finnish Institute for Health and Welfare (THL), Helsinki, Finland | Clinical Groups | Cardiometabolic Diseases Group |

| | | | |
|---|---|---|---|
| **Kaj Metsärinne** | Wellbeing Services County of Southwest Finland, Turku, Finland | Clinical Groups | Cardiometabolic Diseases Group |
| **Jenni Aittokallio** | Wellbeing Services County of Southwest Finland, Turku, Finland | Clinical Groups | Cardiometabolic Diseases Group |
| **Mika Kähönen** | Wellbeing Services County of Pirkanmaa, Tampere, Finland | Clinical Groups | Cardiometabolic Diseases Group |
| **Jussi Hernesniemi** | Wellbeing Services County of Pirkanmaa, Tampere, Finland | Clinical Groups | Cardiometabolic Diseases Group |
| **Daniel Gordin** | Hospital District of Helsinki and Uusimaa, Helsinki, Finland | Clinical Groups | Cardiometabolic Diseases Group |
| **Juha Sinisalo** | Hospital District of Helsinki and Uusimaa, Helsinki, Finland | Clinical Groups | Cardiometabolic Diseases Group |
| **Marja-Riitta Taskinen** | Hospital District of Helsinki and Uusimaa, Helsinki, Finland | Clinical Groups | Cardiometabolic Diseases Group |
| **Tiinamaija Tuomi** | Hospital District of Helsinki and Uusimaa, Helsinki, Finland | Clinical Groups | Cardiometabolic Diseases Group |
| **Timo Hiltunen** | Hospital District of Helsinki and Uusimaa, Helsinki, Finland | Clinical Groups | Cardiometabolic Diseases Group |
| **Jari Laukkanen** | Central Finland Biobank / University of Jyväskylä / Wellbeing Services County of Central Finland, Jyväskylä, Finland | Clinical Groups | Cardiometabolic Diseases Group |
| **Amanda Elliott** | Institute for Molecular Medicine Finland (FIMM), HiLIFE, University of Helsinki, Helsinki, Finland; Broad Institute, Cambridge, MA, USA and Massachusetts General Hospital, Boston, MA, USA | Clinical Groups | Cardiometabolic Diseases Group |
| **Mary Pat Reeve** | Institute for Molecular Medicine Finland (FIMM), HiLIFE, University of Helsinki, Helsinki, Finland | Clinical Groups | Cardiometabolic Diseases Group |
| **Sanni Ruotsalainen** | Institute for Molecular Medicine Finland (FIMM), HiLIFE, University of Helsinki, Helsinki, Finland | Clinical Groups | Cardiometabolic Diseases Group |
| **Dirk Paul** | Astra Zeneca, Cambridge, United Kingdom | Clinical Groups | Cardiometabolic Diseases Group |
| **Natalie Bowers** | Genentech, San Francisco, CA, United States | Clinical Groups | Cardiometabolic Diseases Group |
| **Rion Pendergrass** | Genentech, San Francisco, CA, United States | Clinical Groups | Cardiometabolic Diseases Group |
| **Audrey Chu** | GlaxoSmithKline, Brentford, United Kingdom | Clinical Groups | Cardiometabolic Diseases Group |
| **Kirsi Auro** | GlaxoSmithKline, Espoo, Finland | Clinical Groups | Cardiometabolic Diseases Group |
| **Dermot Reilly** | Johnson & Johnson Innovative Medicine, Boston, MA, United States | Clinical Groups | Cardiometabolic Diseases Group |
| **Mike Mendelson** | Novartis, Boston, MA, United States | Clinical Groups | Cardiometabolic Diseases Group |
| **Jaakko Parkkinen** | Pfizer, New York, NY, United States | Clinical Groups | Cardiometabolic Diseases Group |
| **Melissa Miller** | Pfizer, New York, NY, United States | Clinical Groups | Cardiometabolic Diseases Group |
| **Tuomo Meretoja** | Helsinki University Hospital and University of Helsinki, Helsinki, Finland | Clinical Groups | Oncology Group |
| **Heikki Joensuu** | Helsinki University Hospital and University of Helsinki, Helsinki, Finland | Clinical Groups | Oncology Group |
| **Olli Carpén** | Hospital District of Helsinki and Uusimaa, Helsinki, Finland | Clinical Groups | Oncology Group |
| **Johanna Mattson** | Hospital District of Helsinki and Uusimaa, Helsinki, Finland | Clinical Groups | Oncology Group |
| **Eveliina Salminen** | Hospital District of Helsinki and Uusimaa, Helsinki, Finland | Clinical Groups | Oncology Group |
| **Annika Auranen** | Wellbeing Services County of Pirkanmaa, Tampere, Finland | Clinical Groups | Oncology Group |

| | | | |
|---|---|---|---|
| **Peeter Karihtala** | Helsinki University Hospital and University of Helsinki, Helsinki, Finland | Clinical Groups | Oncology Group |
| **Päivi Auvinen** | Wellbeing services county of North Savo, Kuopio, Finland | Clinical Groups | Oncology Group |
| **Klaus Elenius** | Wellbeing Services County of Southwest Finland, Turku, Finland | Clinical Groups | Oncology Group |
| **Johanna Schleutker** | Wellbeing Services County of Southwest Finland, Turku, Finland | Clinical Groups | Oncology Group |
| **Esa Pitkänen** | Institute for Molecular Medicine Finland (FIMM), HiLIFE, University of Helsinki, Helsinki, Finland | Clinical Groups | Oncology Group |
| **Nina Mars** | Institute for Molecular Medicine Finland (FIMM), HiLIFE, University of Helsinki, Helsinki, Finland | Clinical Groups | Oncology Group |
| **Mark Daly** | Institute for Molecular Medicine Finland (FIMM), HiLIFE, University of Helsinki, Helsinki, Finland; Broad Institute of MIT and Harvard; Massachusetts General Hospital | Clinical Groups | Oncology Group |
| **Relja Popovic** | Abbvie, Chicago, IL, United States | Clinical Groups | Oncology Group |
| **Jeffrey Waring** | Abbvie, Chicago, IL, United States | Clinical Groups | Oncology Group |
| **Bridget Riley-Gillis** | Abbvie, Chicago, IL, United States | Clinical Groups | Oncology Group |
| **Anne Lehtonen** | Abbvie, Chicago, IL, United States | Clinical Groups | Oncology Group |
| **Margarete Fabre** | AstraZeneca, Cambridge, United Kingdom | Clinical Groups | Oncology Group |
| **Jennifer Schutzman** | Genentech, San Francisco, CA, United States | Clinical Groups | Oncology Group |
| **Natalie Bowers** | Genentech, San Francisco, CA, United States | Clinical Groups | Oncology Group |
| **Rion Pendergrass** | Genentech, San Francisco, CA, United States | Clinical Groups | Oncology Group |
| **Diptee Kulkarni** | GlaxoSmithKline, Brentford, United Kingdom | Clinical Groups | Oncology Group |
| **Kirsi Auro** | GlaxoSmithKline, Espoo, Finland | Clinical Groups | Oncology Group |
| **Alessandro Porello** | Johnson & Johnson Innovative Medicine, Spring House, PA, United States | Clinical Groups | Oncology Group |
| **Andrey Loboda** | Merck, Kenilworth, NJ, United States | Clinical Groups | Oncology Group |
| **Heli Lehtonen** | Pfizer, New York, NY, United States | Clinical Groups | Oncology Group |
| **Stefan McDonough** | Pfizer, New York, NY, United States | Clinical Groups | Oncology Group |
| **Sauli Vuoti** | Janssen-Cilag Oy, Espoo, Finland | Clinical Groups | Oncology Group |
| **Kai Kaarniranta** | Wellbeing services county of North Savo, Kuopio, Finland; University of Lodz, Lodz, Poland | Clinical Groups | Opthalmology Group |
| **Joni A Turunen** | Helsinki University Hospital and University of Helsinki, Helsinki, Finland; Folkhälsan Research Center, Helsinki, Finland | Clinical Groups | Opthalmology Group |
| **Terhi Ollila** | Hospital District of Helsinki and Uusimaa, Helsinki, Finland | Clinical Groups | Opthalmology Group |
| **Hannu Uusitalo** | Wellbeing Services County of Pirkanmaa, Tampere, Finland | Clinical Groups | Opthalmology Group |
| **Juha Karjalainen** | Institute for Molecular Medicine Finland (FIMM), HiLIFE, University of Helsinki, Helsinki, Finland | Clinical Groups | Opthalmology Group |
| **Esa Pitkänen** | Institute for Molecular Medicine Finland (FIMM), HiLIFE, University of Helsinki, Helsinki, Finland | Clinical Groups | Opthalmology Group |
| **Mengzhen Liu** | Abbvie, Chicago, IL, United States | Clinical Groups | Opthalmology Group |
| **Erich Strauss** | Genentech, San Francisco, CA, United States | Clinical Groups | Opthalmology Group |
| **Natalie Bowers** | Genentech, San Francisco, CA, United States | Clinical Groups | Opthalmology Group |
| **Hao Chen** | Genentech, San Francisco, CA, United States | Clinical Groups | Opthalmology Group |
| **Rion Pendergrass** | Genentech, San Francisco, CA, United States | Clinical Groups | Opthalmology Group |

| | | | |
|---|---|---|---|
| **Kaisa Tasanen** | Wellbeing services county of North Ostrobothnia, Oulu, Finland | Clinical Groups | Dermatology Group |
| **Laura Huilaja** | Wellbeing services county of North Ostrobothnia, Oulu, Finland | Clinical Groups | Dermatology Group |
| **Katariina Hannula-Jouppi** | Hospital District of Helsinki and Uusimaa, Helsinki, Finland | Clinical Groups | Dermatology Group |
| **Teea Salmi** | Wellbeing Services County of Pirkanmaa, Tampere, Finland | Clinical Groups | Dermatology Group |
| **Sirkku Peltonen** | Wellbeing Services County of Southwest Finland, Turku, Finland | Clinical Groups | Dermatology Group |
| **Leena Koulu** | Wellbeing Services County of Southwest Finland, Turku, Finland | Clinical Groups | Dermatology Group |
| **Nizar Smaoui** | Abbvie, Chicago, IL, United States | Clinical Groups | Dermatology Group |
| **Fedik Rahimov** | Abbvie, Chicago, IL, United States | Clinical Groups | Dermatology Group |
| **Anne Lehtonen** | Abbvie, Chicago, IL, United States | Clinical Groups | Dermatology Group |
| **David Choy** | Genentech, San Francisco, CA, United States | Clinical Groups | Dermatology Group |
| **Rion Pendergrass** | Genentech, San Francisco, CA, United States | Clinical Groups | Dermatology Group |
| **Dawn Waterworth** | Johnson & Johnson Innovative Medicine, Spring House, PA, United States | Clinical Groups | Dermatology Group |
| **Kirsi Kalpala** | Pfizer, New York, NY, United States | Clinical Groups | Dermatology Group |
| **Ying Wu** | Pfizer, New York, NY, United States | Clinical Groups | Dermatology Group |
| **Pirkko Pussinen** | Hospital District of Helsinki and Uusimaa, Helsinki, Finland | Clinical Groups | Odontology Group |
| **Aino Salminen** | Hospital District of Helsinki and Uusimaa, Helsinki, Finland | Clinical Groups | Odontology Group |
| **Tuula Salo** | Hospital District of Helsinki and Uusimaa, Helsinki, Finland | Clinical Groups | Odontology Group |
| **David Rice** | Hospital District of Helsinki and Uusimaa, Helsinki, Finland | Clinical Groups | Odontology Group |
| **Pekka Nieminen** | Hospital District of Helsinki and Uusimaa, Helsinki, Finland | Clinical Groups | Odontology Group |
| **Ulla Palotie** | Hospital District of Helsinki and Uusimaa, Helsinki, Finland | Clinical Groups | Odontology Group |
| **Maria Siponen** | Wellbeing services county of North Savo, Kuopio, Finland | Clinical Groups | Odontology Group |
| **Liisa Suominen** | Wellbeing services county of North Savo, Kuopio, Finland | Clinical Groups | Odontology Group |
| **Päivi Mäntylä** | Wellbeing services county of North Savo, Kuopio, Finland | Clinical Groups | Odontology Group |
| **Ulvi Gursoy** | Wellbeing Services County of Southwest Finland, Turku, Finland | Clinical Groups | Odontology Group |
| **Vuokko Anttonen** | Wellbeing services county of North Ostrobothnia, Oulu, Finland | Clinical Groups | Odontology Group |
| **Kirsi Sipilä** | Oulu University Hospital and University of Oulu, Oulu, Finland | Clinical Groups | Odontology Group |
| **Rion Pendergrass** | Genentech, San Francisco, CA, United States | Clinical Groups | Odontology Group |
| **Hannele Laivuori** | Institute for Molecular Medicine Finland (FIMM), HiLIFE, University of Helsinki, Helsinki, Finland | Clinical Groups | Women's Health and Reproduction Group |
| **Venla Kurra** | Wellbeing Services County of Pirkanmaa, Tampere, Finland | Clinical Groups | Women's Health and Reproduction Group |
| **Laura Kotaniemi-Talonen** | Wellbeing Services County of Pirkanmaa, Tampere, Finland | Clinical Groups | Women's Health and Reproduction Group |
| **Oskari Heikinheimo** | Hospital District of Helsinki and Uusimaa, Helsinki, Finland | Clinical Groups | Women's Health and Reproduction Group |
| **Ilkka Kalliala** | Hospital District of Helsinki and Uusimaa, Helsinki, Finland | Clinical Groups | Women's Health and Reproduction Group |
| **Lauri Aaltonen** | Hospital District of Helsinki and Uusimaa, Helsinki, Finland | Clinical Groups | Women's Health and Reproduction Group |
| **Varpu Jokimaa** | Wellbeing Services County of Southwest Finland, Turku, Finland | Clinical Groups | Women's Health and Reproduction Group |
| **Johannes Kettunen** | Wellbeing services county of North Ostrobothnia, Oulu, Finland | Clinical Groups | Women's Health and Reproduction Group |
| **Marja Vääräsmäki** | Wellbeing services county of North Ostrobothnia, Oulu, Finland | Clinical Groups | Women's Health and Reproduction Group |

| | | | |
|---|---|---|---|
| **Outi Uimari** | Wellbeing services county of North Ostrobothnia, Oulu, Finland | Clinical Groups | Women's Health and Reproduction Group |
| **Laure Morin-Papunen** | Wellbeing services county of North Ostrobothnia, Oulu, Finland | Clinical Groups | Women's Health and Reproduction Group |
| **Maarit Niinimäki** | Wellbeing services county of North Ostrobothnia, Oulu, Finland | Clinical Groups | Women's Health and Reproduction Group |
| **Terhi Piltonen** | Wellbeing services county of North Ostrobothnia, Oulu, Finland | Clinical Groups | Women's Health and Reproduction Group |
| **Katja Kivinen** | Institute for Molecular Medicine Finland (FIMM), HiLIFE, University of Helsinki, Helsinki, Finland | Clinical Groups | Women's Health and Reproduction Group |
| **Elisabeth Widen** | Institute for Molecular Medicine Finland (FIMM), HiLIFE, University of Helsinki, Helsinki, Finland | Clinical Groups | Women's Health and Reproduction Group |
| **Taru Tukiainen** | Institute for Molecular Medicine Finland (FIMM), HiLIFE, University of Helsinki, Helsinki, Finland | Clinical Groups | Women's Health and Reproduction Group |
| **Mary Pat Reeve** | Institute for Molecular Medicine Finland (FIMM), HiLIFE, University of Helsinki, Helsinki, Finland | Clinical Groups | Women's Health and Reproduction Group |
| **Mark Daly** | Institute for Molecular Medicine Finland (FIMM), HiLIFE, University of Helsinki, Helsinki, Finland; Broad Institute of MIT and Harvard; Massachusetts General Hospital | Clinical Groups | Women's Health and Reproduction Group |
| **Niko Välimäki** | University of Helsinki, Helsinki, Finland | Clinical Groups | Women's Health and Reproduction Group |
| **Eija Laakkonen** | University of Jyväskylä, Jyväskylä, Finland | Clinical Groups | Women's Health and Reproduction Group |
| **Jaakko Tyrmi** | University of Oulu, Oulu, Finland / University of Tampere, Tampere, Finland | Clinical Groups | Women's Health and Reproduction Group |
| **Heidi Silven** | University of Oulu, Oulu, Finland | Clinical Groups | Women's Health and Reproduction Group |
| **Eeva Sliz** | University of Oulu, Oulu, Finland | Clinical Groups | Women's Health and Reproduction Group |
| **Riikka Arffman** | University of Oulu, Oulu, Finland | Clinical Groups | Women's Health and Reproduction Group |
| **Susanna Savukoski** | University of Oulu, Oulu, Finland | Clinical Groups | Women's Health and Reproduction Group |
| **Triin Laisk** | Estonian biobank, Tartu, Estonia | Clinical Groups | Women's Health and Reproduction Group |
| **Natalia Pujol** | Estonian biobank, Tartu, Estonia | Clinical Groups | Women's Health and Reproduction Group |
| **Mengzhen Liu** | Abbvie, Chicago, IL, United States | Clinical Groups | Women's Health and Reproduction Group |
| **Bridget Riley-Gillis** | Abbvie, Chicago, IL, United States | Clinical Groups | Women's Health and Reproduction Group |
| **Rion Pendergrass** | Genentech, San Francisco, CA, United States | Clinical Groups | Women's Health and Reproduction Group |
| **Janet Kumar** | GlaxoSmithKline, Collegeville, PA, United States | Clinical Groups | Women's Health and Reproduction Group |
| **Kirsi Auro** | GlaxoSmithKline, Espoo, Finland | Clinical Groups | Women's Health and Reproduction Group |
| **Iiris Hovatta** | University of Helsinki, Finland | Clinical Groups | Depression group |
| **Erkki Isometsä** | Hospital District of Helsinki and Uusimaa, Helsinki, Finland | Clinical Groups | Depression group |
| **Hanna Ollila** | Institute for Molecular Medicine Finland (FIMM), HiLIFE, University of Helsinki, Helsinki, Finland | Clinical Groups | Depression group |
| **Jaana Suvisaari** | Finnish Institute for Health and Welfare (THL), Helsinki, Finland | Clinical Groups | Depression group |
| **Antti Mäkitie** | University of Helsinki and Helsinki University Hospital, Helsinki, Finland | Clinical Groups | ENT (ear, nose and throath) Group |
| **Argyro Bizaki-Vallaskangas** | Wellbeing Services County of Pirkanmaa, Tampere, Finland | Clinical Groups | ENT (ear, nose and throath) Group |
| **Sanna Toppila-Salmi** | University of Eastern Finland and Kuopio University Hospital, Kuopio, | Clinical Groups | ENT (ear, nose and throath) Group |

| | | | |
|---|---|---|---|
| | Finland; Helsinki University Hospital and University of Helsinki, Finland | | |
| **Tytti Willberg** | Wellbeing Services County of Southwest Finland, Turku, Finland | Clinical Groups | ENT (ear, nose and throath) Group |
| **Elmo Saarentaus** | Institute for Molecular Medicine Finland (FIMM), HiLIFE, University of Helsinki, Helsinki, Finland | Clinical Groups | ENT (ear, nose and throath) Group |
| **Antti Aarnisalo** | Hospital District of Helsinki and Uusimaa, Helsinki, Finland | Clinical Groups | ENT (ear, nose and throath) Group |
| **Eveliina Salminen** | Hospital District of Helsinki and Uusimaa, Helsinki, Finland | Clinical Groups | ENT (ear, nose and throath) Group |
| **Elisa Rahikkala** | Northern Ostrobothnia Hospital District, Oulu, Finland | Clinical Groups | ENT (ear, nose and throath) Group |
| **Johannes Kettunen** | Northern Ostrobothnia Hospital District, Oulu, Finland | Clinical Groups | ENT (ear, nose and throath) Group |
| **Kristiina Aittomäki** | Helsinki University Central Hospital, Helsinki, Finland | Clinical Groups | POI (premature ovarian failure) Group |
| **Fredrik Åberg** | Helsinki University Hospital and University of Helsinki, Helsinki, Finland | Clinical Groups | LiverScore Group |
| **Lila Kallio** | Auria Biobank / University of Turku / Wellbeing Services County of Southwest Finland, Turku, Finland | Biobank directors | Biobank directors |
| **Tiina Wahlfors** | THL Biobank / Finnish Institute for Health and Welfare (THL), Helsinki, Finland | Biobank directors | Biobank directors |
| **Jukka Partanen** | Finnish Red Cross Blood Service / Finnish Hematology Registry and Clinical Biobank, Helsinki, Finland | Biobank directors | Biobank directors |
| **Eero Punkka** | Helsinki Biobank / Helsinki University and Hospital District of Helsinki and Uusimaa, Helsinki | Biobank directors | Biobank directors |
| **Raisa Serpi** | Northern Finland Biobank Borealis / University of Oulu / Wellbeing services county of North Ostrobothnia, Oulu, Finland | Biobank directors | Biobank directors |
| **Sanna Siltanen** | Finnish Clinical Biobank Tampere / University of Tampere / Wellbeing Services County of Pirkanmaa, Tampere, Finland | Biobank directors | Biobank directors |
| **Veli-Matti Kosma** | Biobank of Eastern Finland / University of Eastern Finland / Wellbeing services county of North Savo, Kuopio, Finland | Biobank directors | Biobank directors |
| **Tiina Jokela** | Central Finland Biobank / University of Jyväskylä / Wellbeing Services County of Central Finland, Jyväskylä, Finland | Biobank directors | Biobank directors |
| **Anu Jalanko** | Institute for Molecular Medicine Finland (FIMM), HiLIFE, University of Helsinki, Helsinki, Finland | FinnGen Teams | Administration |
| **Auli Toivola** | Institute for Molecular Medicine Finland (FIMM), HiLIFE, University of Helsinki, Helsinki, Finland | FinnGen Teams | Administration |
| **Risto Kajanne** | Institute for Molecular Medicine Finland (FIMM), HiLIFE, University of Helsinki, Helsinki, Finland | FinnGen Teams | Administration |
| **Rodos Rodosthenous** | Institute for Molecular Medicine Finland (FIMM), HiLIFE, University of Helsinki, Helsinki, Finland | FinnGen Teams | Administration |
| **Mervi Aavikko** | Institute for Molecular Medicine Finland (FIMM), HiLIFE, University of Helsinki, Helsinki, Finland | FinnGen Teams | Administration |
| **Helen Cooper** | Institute for Molecular Medicine Finland (FIMM), HiLIFE, University of Helsinki, Helsinki, Finland | FinnGen Teams | Administration |
| **Denise Öller** | Institute for Molecular Medicine Finland (FIMM), HiLIFE, University of Helsinki, Helsinki, Finland | FinnGen Teams | Administration |

| | | | |
|---|---|---|---|
| **Tarja Laitinen** | Institute for Molecular Medicine Finland (FIMM), HiLIFE, University of Helsinki, Helsinki, Finland | FinnGen Teams | Administration |
| **Sofia Kuitunen** | University of Helsinki, Helsinki, Finland | FinnGen Teams | Administration |
| **Mitja Kurki** | Institute for Molecular Medicine Finland (FIMM), HiLIFE, University of Helsinki, Helsinki, Finland; Broad Institute, Cambridge, MA, United States | FinnGen Teams | Analysis |
| **Juha Karjalainen** | Institute for Molecular Medicine Finland (FIMM), HiLIFE, University of Helsinki, Helsinki, Finland | FinnGen Teams | Analysis |
| **Pietro Della Briotta Parolo** | Institute for Molecular Medicine Finland (FIMM), HiLIFE, University of Helsinki, Helsinki, Finland | FinnGen Teams | Analysis |
| **Arto Lehisto** | Institute for Molecular Medicine Finland (FIMM), HiLIFE, University of Helsinki, Helsinki, Finland | FinnGen Teams | Analysis |
| **Juha Mehtonen** | Institute for Molecular Medicine Finland (FIMM), HiLIFE, University of Helsinki, Helsinki, Finland | FinnGen Teams | Analysis |
| **Wei Zhou** | Broad Institute, Cambridge, MA, United States | FinnGen Teams | Analysis |
| **Masahiro Kanai** | Broad Institute, Cambridge, MA, United States | FinnGen Teams | Analysis |
| **Mutaamba Maasha** | Broad Institute, Cambridge, MA, United States | FinnGen Teams | Analysis |
| **Sanni Ruotsalainen** | Institute for Molecular Medicine Finland (FIMM), HiLIFE, University of Helsinki, Helsinki, Finland | FinnGen Teams | Analysis |
| **Susanna Lemmelä** | Institute for Molecular Medicine Finland (FIMM), HiLIFE, University of Helsinki, Helsinki, Finland | FinnGen Teams | Analysis |
| **Aki Havulinna** | Institute for Molecular Medicine Finland (FIMM), HiLIFE, University of Helsinki, Helsinki, Finland; Finnish Institute for Health and Welfare (THL), Helsinki, Finland | FinnGen Teams | Clinical Endpoint Development |
| **L. Elisa Lahtela** | Institute for Molecular Medicine Finland (FIMM), HiLIFE, University of Helsinki, Helsinki, Finland | FinnGen Teams | Clinical Endpoint Development |
| **Mari Kaunisto** | Institute for Molecular Medicine Finland (FIMM), HiLIFE, University of Helsinki, Helsinki, Finland | FinnGen Teams | Communication |
| **Elina Kilpeläinen** | Institute for Molecular Medicine Finland (FIMM), HiLIFE, University of Helsinki, Helsinki, Finland | FinnGen Teams | E-Science |
| **Tianduanyi Wang** | Institute for Molecular Medicine Finland (FIMM), HiLIFE, University of Helsinki, Helsinki, Finland | FinnGen Teams | E-Science |
| **Timo P. Sipilä** | Institute for Molecular Medicine Finland (FIMM), HiLIFE, University of Helsinki, Helsinki, Finland | FinnGen Teams | E-Science |
| **Oluwaseun Alexander Dada** | Institute for Molecular Medicine Finland (FIMM), HiLIFE, University of Helsinki, Helsinki, Finland | FinnGen Teams | E-Science |
| **Awaisa Ghazal** | Institute for Molecular Medicine Finland (FIMM), HiLIFE, University of Helsinki, Helsinki, Finland | FinnGen Teams | E-Science |
| **Rigbe Weldatsadik** | Institute for Molecular Medicine Finland (FIMM), HiLIFE, University of Helsinki, Helsinki, Finland | FinnGen Teams | E-Science |
| **Sanni Ruotsalainen** | Institute for Molecular Medicine Finland (FIMM), HiLIFE, University of Helsinki, Helsinki, Finland | FinnGen Teams | E-Science |
| **Jaska Uimonen** | Institute for Molecular Medicine Finland (FIMM), HiLIFE, University of Helsinki, Helsinki, Finland | FinnGen Teams | E-Science |

| | | | |
|---|---|---|---|
| **Kati Donner** | Institute for Molecular Medicine Finland (FIMM), HiLIFE, University of Helsinki, Helsinki, Finland | FinnGen Teams | Genotyping |
| **Anu Loukola** | Helsinki Biobank / Helsinki University and Hospital District of Helsinki and Uusimaa, Helsinki | FinnGen Teams | Sample Collection Coordination |
| **Päivi Laiho** | THL Biobank / Finnish Institute for Health and Welfare (THL), Helsinki, Finland | FinnGen Teams | Sample Logistics |
| **Susanna Lemmelä** | Institute for Molecular Medicine Finland (FIMM), HiLIFE, University of Helsinki, Helsinki, Finland | FinnGen Teams | Registry Data Operations |
| **Sami Koskelainen** | THL Biobank / Finnish Institute for Health and Welfare (THL), Helsinki, Finland | FinnGen Teams | Registry Data Operations |
| **Tero Hiekkalinna** | THL Biobank / Finnish Institute for Health and Welfare (THL), Helsinki, Finland | FinnGen Teams | Registry Data Operations |
| **Teemu Paajanen** | THL Biobank / Finnish Institute for Health and Welfare (THL), Helsinki, Finland | FinnGen Teams | Registry Data Operations |
| **Arto Pietilä** | THL Biobank / Finnish Institute for Health and Welfare (THL), Helsinki, Finland | FinnGen Teams | Registry Data Operations |
| **Priit Palta** | Institute for Molecular Medicine Finland (FIMM), HiLIFE, University of Helsinki, Helsinki, Finland | FinnGen Teams | Sequencing Informatics |
| **Mary Pat Reeve** | Institute for Molecular Medicine Finland (FIMM), HiLIFE, University of Helsinki, Helsinki, Finland | FinnGen Teams | Phenotype team |
| **Shanmukha Sampath Padmanabhuni** | Institute for Molecular Medicine Finland (FIMM), HiLIFE, University of Helsinki, Helsinki, Finland | FinnGen Teams | Phenotype team |
| **Harri Siirtola** | University of Tampere, Tampere, Finland | FinnGen Teams | Phenotype team |
| **Javier Gracia-Tabuenca** | University of Tampere, Tampere, Finland | FinnGen Teams | Phenotype team |
| **Marika Kaakinen** | Institute for Molecular Medicine Finland (FIMM), HiLIFE, University of Helsinki, Helsinki, Finland | FinnGen Teams | Phenotype team |
| **Shuang Luo** | Institute for Molecular Medicine Finland (FIMM), HiLIFE, University of Helsinki, Helsinki, Finland | FinnGen Teams | Phenotype team |
| **Vincent Llorens** | Institute for Molecular Medicine Finland (FIMM), HiLIFE, University of Helsinki, Helsinki, Finland | FinnGen Teams | Phenotype team |
| **Iina Laak** | Institute for Molecular Medicine Finland (FIMM), HiLIFE, University of Helsinki, Helsinki, Finland | FinnGen Teams | Data protection officer |
| **Johanna Mäkelä** | Finnish Biobank Cooperative – FINBB | FinnGen Teams | FINBB - Finnish biobank cooperative |
| **Pauli Wihuri** | Finnish Biobank Cooperative – FINBB | FinnGen Teams | FINBB - Finnish biobank cooperative |
| **Tom Southerington** | Finnish Biobank Cooperative – FINBB | FinnGen Teams | FINBB - Finnish biobank cooperative |
| **Meri Lähteenmäki** | Finnish Biobank Cooperative – FINBB | FinnGen Teams | FINBB - Finnish biobank cooperative |
